# Longitudinal analysis of genetic and environmental interplay in human metabolic profiles and the implication for metabolic health

**DOI:** 10.1101/2024.09.23.24314199

**Authors:** Jing Wang, Alberto Zenere, Xingyue Wang, Göran Bergström, Fredrik Edfors, Mathias Uhlén, Wen Zhong

## Abstract

**Background:** Understanding how genetics and environmental factors shape human metabolic profiles is crucial for advancing metabolic health. Variability in metabolic profiles, influenced by genetic makeup, lifestyle, and environmental exposures, plays a critical role in disease susceptibility and progression.

**Methods:** We conducted a two-year longitudinal study involving 101 clinically healthy individuals aged 50 to 65, integrating genomics, metabolomics, lipidomics, proteomics, clinical measurements, and lifestyle questionnaire data from repeat sampling. We evaluated the influence of both external and internal factors, including genetic predispositions, lifestyle factors, and physiological conditions, on individual metabolic profiles. Additionally, we developed an integrative metabolite-protein network to analyze protein-metabolite associations under both genetic and environmental regulations.

**Results:** Our findings highlighted the significant role of genetics in determining metabolic variability, identifying 22 plasma metabolites as genetically predetermined. Environmental factors such as seasonal variation, weight management, smoking, and stress also significantly influenced metabolite levels. The integrative metabolite-protein network comprised 5,649 significant protein-metabolite pairs and identified 87 causal metabolite-protein associations under genetic regulation, validated by showing a high replication rate in an independent cohort. This network revealed stable and unique protein-metabolite profiles for each individual, emphasizing metabolic individuality. Notably, our results demonstrated the importance of plasma proteins in capturing individualized metabolic variabilities. Key proteins representing individual metabolic profiles were identified and validated in the UK Biobank, showing great potential for predicting metabolic diseases and metabolic risk assessment.

**Conclusions:** Our study provides longitudinal insights into how genetic and environmental factors shape human metabolic profiles, revealing unique and stable individual metabolic profiles. Plasma proteins emerged as key indicators for capturing the variability in human metabolism and assessing metabolic risks. These findings offer valuable tools for personalized medicine and the development of diagnostics for metabolic diseases.

## Background

The human metabolome is dynamic, and the variability in human metabolic profiles across individuals is shaped by each person’s unique genetic makeup, lifestyle, and environmental exposures^1,2^. These factors play critical roles in disease susceptibility and progression, including obesity, diabetes, hypertension, cardiovascular disease and other metabolic abnormalities^3–6^. Despite significant advancements in metabolomics, the determinants of individual metabolic variability remain incompletely understood. Twin studies have revealed a broad range of heritability for metabolite levels in human plasma^7,8^, and genome-wide association studies (GWAS) have identified numerous genetic variants influencing metabolite levels (mQTL, metabolite quantitative trait loci)^9–13^. This demonstrates the important role of genetics in human metabolism for the understanding of individual metabolic diversity. Beyond genetics, human metabolic profiles are also influenced by various factors such as obesity^14–16^, lifestyle ^17^, diet^2,18,19^, microbiome^2,18,19^, medications^20^, and other environmental exposures. For instance, by analyzing fasting plasma profiles of 1,183 metabolites in 1,679 samples from 1,368 individuals, Chen et al.^21^ found that diet, genetics and the gut microbiome could explain 9.3%, 3.3% and 12.8%, respectively, of the inter-individual variations in plasma metabolomics.

However, these GWAS and association studies often overlook the temporal dynamics and environmental interactions that continuously influence the metabolome. Seasonal variations, for instance, introduce another layer of complexity in metabolite levels^22–24^, reflecting changes in environmental conditions, dietary habits, physical activity, and other lifestyle factors^25,26^. The heterogeneity in metabolite levels among individuals further underscores their multifaceted roles in various biological processes^27,28^. To accurately contextualize this variability in human metabolism, incorporating longitudinal molecular data from the same individuals is crucial. This approach allows for the monitoring of dynamics in metabolite levels in response to external influences and internal physiological changes, providing a more comprehensive understanding of individual metabolite variability^5,29,30^. In addition, longitudinal data can reveal temporal metabolic regulation patterns that are not apparent in cross-sectional studies^31^, offering deeper insights into the dynamic interplay between genetic predispositions and environmental factors and advancing precision medicine for metabolic health.

Here, we conducted a detailed longitudinal multi-omics analysis involving 101 individuals aged 50-65 over two years to explore the dynamics and individual differences in metabolic profiles. The influences of genetic predispositions, lifestyle factors, and physiological conditions on individual metabolic profiles have been investigated. Additionally, we established an integrative metabolite-protein network and identified key proteins and metabolites associated with human metabolic risk. This work contributes to a more comprehensive understanding of individual metabolite variability and advances our knowledge in more personalized approaches to monitoring metabolic health.

## Methods

### The wellness profiling study

The Swedish SciLifeLab SCAPIS Wellness Profiling (S3WP) study is an observational study aimed at gathering longitudinal clinical and molecular data from a community-based cohort. This study derived from the Swedish CArdioPulmonary bioImage Study (SCAPIS), which is a prospective observational study with 30,154 individuals aged 50 to 65 years at enrollment, randomly selected from the general Swedish population between 2015 and 2018 ^32^. In SCAPIS, no exclusion criteria are applied except the inability to understand written and spoken Swedish for informed consent. In the S3WP study, we enrolled 101 healthy individuals with the following exclusion criteria: (1) previously received health care for myocardial infarction, stroke, peripheral artery disease, or diabetes, (2) presence of any clinically significant disease which, in the opinion of the investigator, may interfere with the results or the subject’s ability to participate in the study, (3) any major surgical procedure or trauma within 4 weeks of the first study visit, or (4) medication for hypertension or hyperlipidemia. Before enrolling in the S3WP study, all subjects had been extensively assessed by SCAPIS ^32^. Throughout the duration of the S3WP study, follow-up visits are conducted every third month (±2 weeks) in the first year and approximately a 6-month interval in the second year. All subjects were fasting overnight (at least 8 h) before the visits. Lifestyle questionnaires, anthropometric measurements, clinical measurements, and plasma proteome profiling, metabolome profiling and lipidome profiling were examined at each of the follow-up visit. Whole genome sequencing data were detected at the baseline (**Fig. 1a, Additional file 2: Table S1**). The study has been approved by the Ethical Review Board of Göteborg, Sweden (registration number 407-15), and all participants provided written informed consent. The study protocol adheres to the ethical guidelines outlined in the 1975 Declaration of Helsinki.

**Fig. 1.**
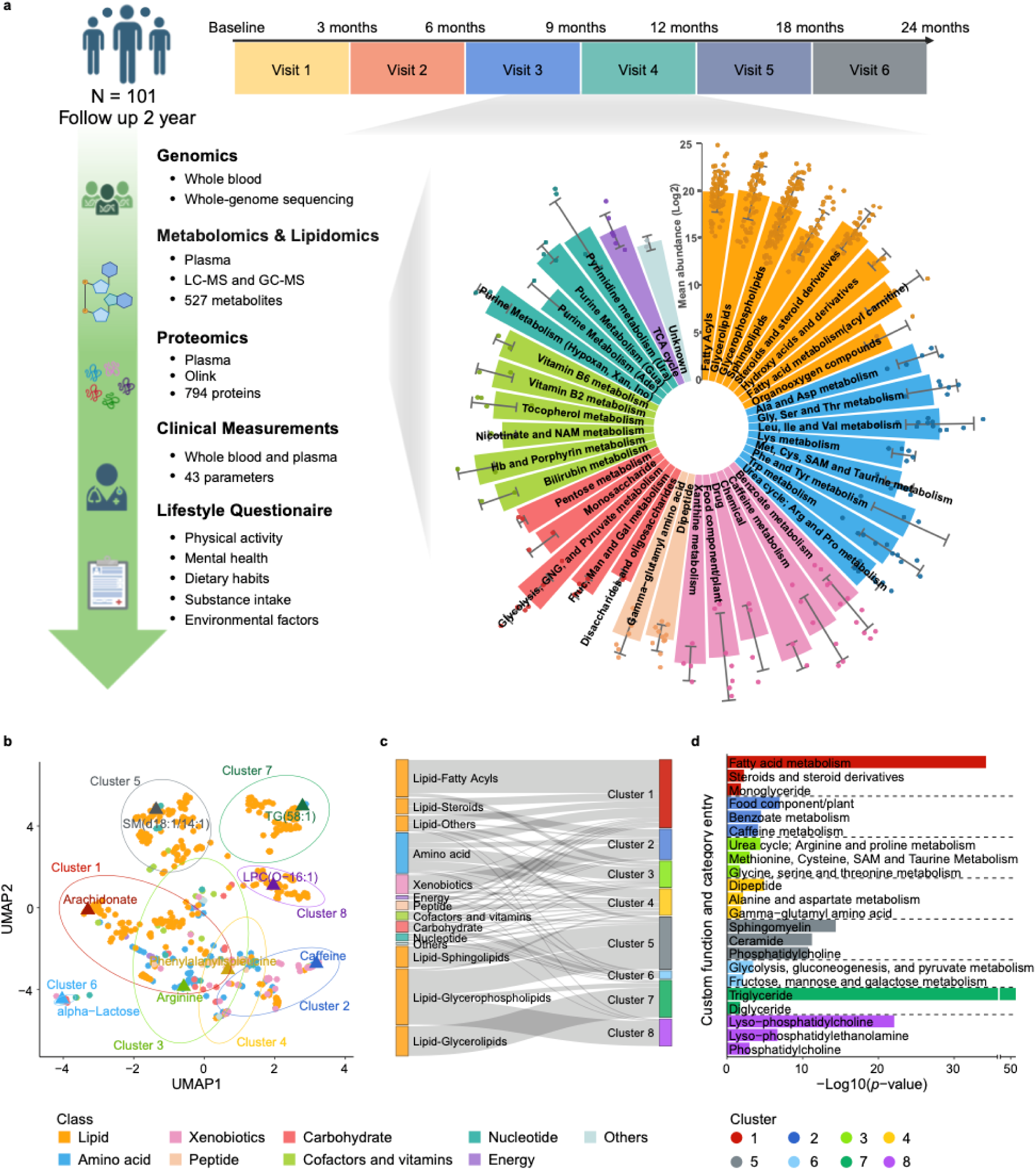
Longitudinal multi-omics profiling and co-expression analysis of human plasma metabolites in 101 healthy individuals. **a** Overview of the study design for the S3WP project, detailing the longitudinal data collection strategy over six visits in two years (created with biorender.com). This included whole-genome sequencing, proteomics, metabolomics, lipidomics, clinical measurements, and lifestyle questionnaires from 101 individuals. A total of 527 identified metabolites were categorized into 9 major classes. **b** UMAP clustering of the 527 metabolites showing their co-expression patterns, color-coded by their classes and grouped by co-expression clusters. **c** Sankey plot showing the distribution of metabolic classes across the clusters. **d** Bar plot showing the functional enrichment results of the metabolite clusters using Fisher’s exact test.

### Self-reported questionnaires

Self-reported questionnaires, comprised 140 questions, were used to gather detailed information covering health, family history, medication, occupational and environmental exposure, lifestyle, psychosocial well-being, socioeconomic status, and other social determinants. These questionnaires had been administered in the SCAPIS trial. During each visit in the S3WP program, a selection of these questions was repeated to update the information from the initial SCAPIS questionnaire. Additionally, participants were asked about any changes in lifestyle factors between visits, such as infections, disease, medication, perceived health, and exercise level.

### Anthropometric and clinical measurements

Height was measured to the nearest centimeter without shoes, with participants wearing indoor clothing. Weight was recorded on a calibrated digital scale under the same conditions. BMI was calculated by dividing weight (kg) by the square of height (m). Waist circumference was measured midway between the iliac crest and the lowest rib margin in the left and right mid-axillary lines. Hip circumference was measured at the widest part of the buttocks. Bioimpedance was assessed using the Tanita MC-780MA following the manufacturer’s instructions. Systolic and diastolic blood pressure were measured in the supine position after a 5-minute rest using the Omron P10 automatic device. Blood pressure was initially measured in both arms during the first visit, and subsequently in the arm that showed the highest reading. Clinical chemistry and hematology assessments included capillary glucose (Hemocue), plasma glucose, HbA1c, triglycerides, total cholesterol, LDL, HDL, ApoA1, ApoB, ApoA1/B ratio, creatinine, high-sensitive C-reactive protein (hsCRP), ALAT, GGT, urate, cystatin C, vitamin D, TNT, NTproBNP, hemoglobin, and blood cell count. The complete list of the clinical variables can be found in **Additional file 2: Table S1**.

### Whole genome sequencing

The whole genome-sequencing procedure has been detailed previously by Zhong et al. ^33,34^. Briefly, Genomic DNA was sequenced to average 30X coverage on the HiSeq X system (Illumina, paired-end 2 × 150 bp). The alignment was performed using BWAmem using GRCh38.p7 as reference genome. Single-nucleotide and insertion/deletion variants were called following the GATK pipeline (https://software.broadinstitute.org/gatk/best-practices; GATK v3.6). BCFtools^35^ and PLINK 2.0^36^ were used to perform quality control (QC). The exclusion criteria included: (1) removing variants which did not receive the “PASS” tag from GATK; (2) removing variants with minQUAL <30; (3) removing variants/samples that with a genotyping rate < 0.05; (5) removing variants with a low minor allele frequency (MAF) (<5%); (6) removing variants that failed the Hardy–Weinberg equilibrium (HWE) test (*P* < 1×10^-6^). In total, 6,691,390 high-quality variants were identified in all samples. Functional annotation of variants was performed using Ensembl Variant Effect Predictor (VEP) v111^37^.

### Plasma metabolomics and lipidomics profiling

Plasma metabolites and lipids profiling were obtained by gas chromatography-mass spectrometry (GC-MS) and liquid chromatography-mass spectrometry (LC-MS)^29^. Briefly, the metabolites were extracted by using Agilent 1290 Infinity UHPLC-system (reverse phase chromatography) combined with an Agilent 6550 Q-TOF mass spectrometer equipped with an electrospray Jetstream ion source operating in positive and negative ion mode. The *m*/*z* range was 70-1700, and data were collected in centroid mode with an acquisition rate of 4 scans/s. The mass spectrometry files were processed using Profinder B.08.01 (Agilent Technologies Inc., Santa Clara, CA, USA). Peak detection was performed using mass feature extraction. The lipids were extracted following a modified Folch protocol ^38^. The data were processed using Batch Targeted Feature Extraction algorithm within MassHunter™ ProFinder version B.08.00 (Agilent Technologies Inc., Santa Clara, CA, USA). In-house databases with exact mass and experimental retention times of lipids were used for identification. The detailed method can be found in Tebani et al.^29^. In total, 456 metabolites were identified in 101 subjects and 173 lipids were measured in 50 subjects.

All metabolite concentrations were log2-transformed to approximate a normal distribution. Metabolites with any of the following conditions have been removed: (1) metabolites that failed detection in at least 30% of samples; (2) the ratio of maximum and minimum interquartile range (IQR) of metabolite concentrations across four visits > 2; (3) duplicated metabolites. In addition, 5 subjects were removed because they participated in < 4 follow-ups. The filtering process retained a total of 527 unique metabolites for 96 subjects (357 metabolites based on 96 subjects and 170 lipids based on 50 subjects) for the downstream analysis of the study.

Metabolite annotation was performed using resources from the Human Metabolomics Database (HMDB) (version 5.0) ^39^ and relevant literature ^4,7–10^. A total of 527 metabolites were categorized into 9 main classes and 63 subclasses. Additionally, 11 lipid subclasses were further subdivided into 23 secondary lipid subclasses, resulting in the creation of 75 custom terms. These customized terms, along with metabolic pathways curated in the Small Molecule Pathway Database (SMPDB), were used for the enrichment analysis of metabolites. The complete list of the customized terms can be found in **Additional file 2: Table S3.**

### Plasma protein profiling

We used a multiplex proximity extension assay (Olink Bioscience, Uppsala Sweden) to measure the relative concentrations of 794 plasma proteins in eleven Olink panels. To minimize inter-run and intra-run variation, the samples were randomized across plates and normalized using both an internal control (extension control) and an inter-plate control; then a pre-determined correction factor was applied to transform the data. The pre-processed data were provided in the arbitrary unit Normalized Protein eXpression (NPX) on a log2 scale. QC procedures were performed at both sample and protein level. Briefly, samples were flagged (did not pass QC) if the incubation control deviated more than a pre-determined value (+/− 0.3) from the median value of all samples on the plate (www.olink.com). To reduce the batch effect between samples run at different times, bridging reference samples from different visits were also run on plates from the different batches. Reference sample normalization based on bridging samples was conducted to minimize technical variation between batches (www.olink.com). After QC, a total of 794 unique proteins for 90 subjects and 6 visits (540 samples) were retained for analysis. The detailed information about plasma protein profiling can be found in previous papers^34,33^. Proteins were annotated according to their molecular function, following the Human Protein Atlas v23 (https://www.proteinatlas.org/).

### Co-expression analysis of plasma metabolites

Before performing co-expression analysis, metabolomics and lipidomics data for 527 metabolites were scaled to zero mean and unit variance. UMAP was applied as an unsupervised clustering modeling method for dimensionality reduction, providing an overview of clustering patterns among samples with similar data profiles. The K Nearest Neighbour Search (KNN) algorithm was implemented to calculate the adjacency matrix using the ‘nn2’ function from the R package RANN v2.6.1^40^, setting the maximum number of nearest neighbors to 20. To calculate the number of shared nearest neighbors (SNN), we employed the ‘sNN’ function from the R package dbscan v1.1.11^41^, considering 5 neighbors for the calculation. R package igraph v1.5.0^42^ was used to build the adjacency matrix based on the nearest neighbor results for each metabolite and to identify communities using the ‘cluster_louvain’ function. In total, 19 clusters were identified from the initial clustering. The mean scaled abundance of metabolites within each of these 19 clusters was then used for a second step of hierarchical clustering. Euclidean distances between the 19 clusters were computed, and similarities were assessed using the ward.D2 method. The hierarchical clustering dendrogram was subsequently divided into 8 distinct groups, resulting in the identification of 8 unique metabolite clusters (**Additional file 2: Table S2**).

### Seasonal variation analysis

The seasonal patterns of both metabolites and proteins were analyzed by calculating the amplitude of their temporal expressions. The amplitude was defined as the square root of the sum of the squares of the coefficients of the sine and cosine terms of sampling month from the fitted seasonal model:

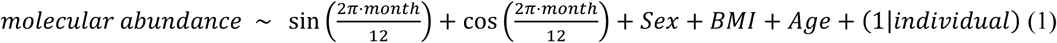

To identify temporally co-expressed metabolites and proteins, Euclidean distances were calculated for those showing significant results associated with sampling month (FDR-adjusted P < 0.05) based on their scaled mean intensity at each month. Clustering analysis was then performed using the ‘ward.D2’ method in the ‘hclust’ function from the R ‘stats’ package. The resulting hierarchical clustering tree was divided into distinct seasonal groups, revealing unique patterns of seasonal variation (**Additional file 2: Table S5 and Additional file 2: Table S8**).

### Genome-wide association analysis of plasma metabolites

For each plasma metabolite, we calculated the coefficients of metabolite intensity for each subject using a linear regression model, with visit included as a covariate. These coefficients served as adjusted metabolite levels for subsequent GWAS analysis. GWAS was conducted using the PLINK v2.0^36^, employing a linear regression model that included body mass index (BMI), sex, and age as covariates. To identify independent mQTLs, linkage disequilibrium (LD) *r*^2^ > 0.1 with window size 1 Mb was first used to exclude the correlated variants. Given the high correlation among metabolites, we utilized an eigendecomposition method to estimate the effective number of independent metabolites^4^. Out of the 527 metabolites, the estimated number of independent metabolites was 23, and this number was used to calculate the Bonferroni threshold for multiple hypothesis testing (5 × 10^−8^ / 23 = 2.17 × 10^−9^). For metabolites associated with multiple mQTLs, conditional analysis was performed by re-calculating genetic associations using the lead single-nucleotide polymorphism (SNP) as a covariate. Only associations with a conditional *P*-value < 0.01 were considered to be independent mQTLs. A total of 66 significant associations between genetic variants and individual blood metabolite concentrations were identified. Among them, 19 independent metabolite quantitative trait loci (mQTLs) (Linkage Disequilibrium, LD, *r*^2^ < 0.1, conditional *P* < 0.01) involving 22 metabolites were identified (**Additional file 2: Table S11**).

### Meta-analysis of plasma metabolite GWAS

For the meta-analysis of plasma metabolite GWAS, we included six out of 22 identified genetic-related metabolites with available published GWAS summary results. The GWAS summary data from published studies^4,7^ was retrieved via the GWAS Catalog (www.ebi.ac.uk/gwas/, accession date: 2023.10), including 3 cohorts: Cooperative Health Research in the Region of Augsburg (KORA, N = 1768), TwinsUK (N = 1052) and Canadian Longitudinal Study of Aging (CLSA, N = 8,299). All these three cohorts comprised relatively healthy European individuals. To ensure consistency across datasets, we used LiftOver^43^ to convert the genome coordinates to the GRCh38 reference genome. The meta-analysis was performed on these three cohorts along with our study, involving a total of 5.8 million SNPs, using an inverse-variance fixed-effect model using GWAMA v2.2.2^44^. The genome-wide significance threshold for the meta-analysis was set at *P* < 8.33×10^-9^, accounting for multiple testing correction (5×10^-8^ / 6).

### Experimental validation

ACADS short interfering RNA (siRNA) and negative control siRNA were transfected into 293T cells. Following a 48-hour incubation period, the cells were collected and divided into two equal portions. One portion was lysed and quantified using the Bicinchoninic Acid (BCA) Protein Assay Kit. Equal amounts of protein lysate from each sample were used for immunoblotting to assess the expression levels of the ACADS protein. The immunoblotting results were analyzed to determine the efficacy of ACADS knockdown. The remaining portion of the cells was processed for metabolite analysis. Specifically, the relative content of butyrylcarnitine was measured using a liquid chromatography-quadrupole time-of-flight mass spectrometer (LC-QTOF MS, Agilent #1290-6546). The mass spectrometer was equipped with Agilent Jet-stream source operating in negative and positive ion mode with source parameters set as follow: Nebulizer gas, 45psi; Sheath gas temperature, 325 °C; Sheath gas flow, 10L/min; Dry gas temperature, 280 °C; Dry gas flow, 8L/min; Capillary voltage, 3500v for two ion modes and nozzle voltage, 500v for positive and 1000v for negative mode. The QTOF scan parameters were set as follows: Scan speed, 2 scan/s. Separation of metabolites was achieved in a Waters ACQUITY UPLC BEH Amide column(2.1 mm × 100 mm × 1.7 μm) and guard column (2.1 mm × 5 mm × 1.7 μm). Agilent Masshunter profinder 10.0 was used to extract its characteristic m/z 232.1547 ion from the total ion chromatogram. Butyrylcarnitine was quantified by peak area. Data analysis was conducted using Agilent MassHunter profinder 10.0 software. The detailed experimental setup are provided in **Supplementary Information**.

### Protein-metabolite association analysis, Mendelian randomization, and network analysis

Linear mixed modeling (LMM) was conducted to identify associations between 527 metabolites and 794 proteins, with sex, age, and BMI as fixed effects, and subject and visit as random effects. The analysis was performed using the ‘lmerTest’ package^45^, and the Kenward-Roger approximation was applied to calculate *P*-value using the R package ‘pbkrtest’ v0.5.2^46^. In addition, one-sample MR analysis was performed to test the causal relationships between protein-metabolite pairs identified through LMM (FDR-adjusted *P* < 0.05). This analysis utilized instrumental variable (IV) regression by two-stage least squares (2SLS) using the ‘ivreg’ function from the R package AER 1.2-10^47^, which is based on Sex, age and BMI were included in the regression models as covariates, while the independent pQTLs (genome-wide significance, *P* < 5×10^-8^) associated with the protein were used as IVs. SNP would be removed from the IVs if it had more than 5 association across the proteins^13^. The full list of IVs was provided in **Additional file 2: Table S15.**

The protein-metabolite association network was constructed by combining both the LMM and MR results. Betweenness centrality score for each node in the network was calculated using the function ‘betweenness’ from the package ‘igraph’ v1.5.0^42^. Proteins and metabolites with a score > MAD above the respective median were classified as Tier 2, while proteins and metabolites with a score >2×MAD the median were categorized as Tier 1. All significant (FDR-adjusted *P* < 0.05) protein-metabolite pairs identified from the LMM were used to perform a UMAP clustering based on samples without missing values.

### Metabolic risk stratification of participants in the study

To evaluate the metabolic risk levels of participants at various study visits, five classical metabolic risk indicators were utilized: body mass index (BMI), high-density lipoprotein (HDL), systolic blood pressure (SBP), fasting glucose (Gluc), and triglycerides (TG). Each measurement was normalized by adjusting for sex-specific effects and then dividing by the standard deviation across all samples. K-means clustering was performed to stratify the samples into two groups (high risk and low risk) using the function ‘kmeans’ from the R package ‘stats’ ^48^. Additionally, other clinical biochemical markers, including alanine aminotransferase (ALAT), gamma-glutamyltransferase (GGT), urate, troponin T (TNT), and white blood cells (WBC), were normalized in a same way. These markers were then used to test for differential expression levels between the two stratified metabolic risk groups.

### Individuals with metabolic diseases in the UK Biobank

The UK Biobank is a large-scale biomedical database and research resource that includes genetic, lifestyle, and health information from half a million UK participants aged 40-69 at baseline^49,50^. Participants were enrolled from 2006 to 2010 in 22 recruitment centers across the UK, with follow-up data continuously collected. Proteomic profiling was conducted on blood plasma samples collected during baseline recruitment from a randomized subset of UK Biobank participants. For this study, we focused on Normalized Protein Expression (NPX) data for Tier 1 proteins and extracted metabolic disease diagnoses information (field ID: 41202, ICD 10: E10, E11, E66, E03, E05, E78, M10, E88). Participants with available proteome data for Tier 1 proteins were selected. To validate the predictive power of Tier 1 proteins for various metabolic diseases, we focused on diseases with sufficient case numbers for proteome analysis. Only data from participants diagnosed before baseline recruitment were included, and individuals with multiple diagnoses were excluded. This resulted in the inclusion of type 2 diabetes (N = 144), obesity (N = 25), hyperthyroidism (N = 21), and gout (N = 27) for predictive analysis.

For robust statistical analysis, NPX values were rank-inverse normal-transformed. Remaining missing values were imputed using the average value calculated across all individuals. Ultimately, the dataset comprised 242 participants in the metabolic disease group and 5,511 in the healthy group.

### Identification of individuals who develop obesity in the UK Biobank

To identify individuals at risk of developing obesity in the future, we utilized BMI records (field ID:21001) from the UK Biobank, which included baseline recruitment data collected between 2006 and 2010 and three follow-up visits in 2012-2013, 2014+, and 2019+. Participants with a baseline BMI lower than 25 kg/m² were included in the analysis. These individuals were further stratified into two groups: the ‘future obese’ group, consisting of individuals who recorded a BMI higher than 30 kg/m² in at least one of the three follow-up visits, and the ‘control’ group, comprising individuals who maintained a BMI lower than 30 kg/m² across all subsequent visits. To ensure data quality and relevance, we filtered the samples based on the availability of proteome data for Tier 1 proteins. This filtering process resulted in the classification of fifteen individuals into the ‘future obesity’ group and 3,185 individuals into the control group.

### Machine learning for predictive tasks

To develop and validate prediction models, the samples were randomly split into training and validation datasets using a 7:3 ratio. For analyses performed on the S3WP data, samples from the same individuals were kept together in either the training or validation group. For the UK Biobank data, balanced datasets were constructed by randomly selecting an equal number of control samples for predicting metabolic diseases. All data were scaled before the prediction analysis. The prediction models were built using the XGBClassifier from the Python library xgboost v2.0.3^51^ with “binary:logistic” used as the objective. To address class imbalance, the subsample parameter was set to 0.5, and the scale_pos_weight parameter was adjusted to the ratio of negative and positive samples. The model training and validation procedure was repeated 100 times, each time with a different random split of the data, to account for stochastic variability in the selection of training and validation data. Receiver operating characteristic (ROC) curves were generated and visualized using the R package ‘verification’ v1.42^52^.

### Statistical analysis and visualization

Uniform Manifold Approximation and Projection (UMAP) was performed by using the R package umap^53^. Canonical correspondence analysis (CCA) was performed including lifestyle factors, anthropometric and clinical measurements, and visit as constraining variables using the R package vegan v2.6.4^54^. The association between each variable and CCA1 and CCA2 was quantified by the estimated coefficient obtained from univariate linear regression. The mixed model expressed by equation (1) was used to determine the associations between sex, BMI, age, seasonality and metabolite abundance, with metabolite abundance as response, sex, BMI, age, and month of sampling as fixed effects and subject as random effect. All of mixed model analyses were performed using the lmerTest package^45^ and Kenward-Roger approximation was applied to calculate *P*-value using the R package pbkrtest v0.5.2^46^. Variance analysis of metabolites and UMAP components was conducted using a linear regression model including significantly associated mQTLs (and pQTLs for UMAP components), clinical measurements, lifestyle parameters, sex, age, and visit as variables. The fraction of explained variability was measured as the Sum of Squares Explained (SSE) and *p*-value was determined using Analysis of Variance (ANOVA) by the built-in R function aov. Pearson correlation was used to estimate the correlation between metabolites and proteins. Kruskal-Walis test and *t*-test were used to compare differences in the levels of metabolites or proteins between groups. Fisher’s exact test was used for enrichment analysis of metabolites. FDR was calculated for multiple testing correction by using the function p.adjust, selecting the Benjamini-Hochberg method. FDR-adjusted *P* <0.05 was considered significant in the analysis.

The Sankey plot was generated using the function sankeyNetwork of R package networkD3 v0.4^55^. The heatmap was generated using the R package pheatmap v1.0.12^56^. Networks were created using the R package igraph v1.5.0^42^. Chord diagrams were generated using the R packages circlize^57^. Spider plots were generated using the package ggradar v0.2^84^. The package ggalluvial v0.12.5^58^ was used to show the group of the samples from the same individual across the six visits. All the other visualizations were performed using the ggplot2^59^ R package. All of the data analysis and visualization was performed using the R project^60^.

## Results

### Longitudinal analysis of human metabolic profiles in a wellness study

To systematically investigate the human metabolic profiles over time, we performed a comprehensive integrative multi-omics analysis of 101 participants in the Swedish SciLifeLab SCAPIS Wellness Profiling (S3WP) program over two years ^29^. Whole-genome sequencing was performed at baseline for all participants. Extensive phenotyping of the individuals was conducted every three months in the first year and at approximately a 6-month interval in the second year, which included plasma metabolome and lipidome profiling, proteome profiling, clinical measurements, and detailed lifestyle questionnaires covering physical activity, mental health, substance intake, and other environmental factors, alongside blood sample collection, to capture seasonal fluctuations and provide a robust temporal perspective (**Fig. 1a and Additional file 2: Table S1**). Using a combination of GC-MS and LC-MS technologies, we identified a total of 527 metabolites in the study. These metabolites were classified into nine main classes, covering a wide range of lipids (n = 335, 63.6%), amino acids (n = 77, 14.6%), xenobiotics (n = 37, 7.0%), peptides (n = 17, 3.2%), carbohydrates (n = 20, 3.8%), cofactors and vitamins (n = 16, 3.0%), nucleotides (n = 14, 2.7%), energy (n =7, 1.3%), and other metabolites (n = 4, 0.8%). The metabolites were further categorized into 63 subclasses based on their functions and biochemical characteristics (**Fig. 1a and Additional file 2: Table S2, see Methods for more details**).

To explore the co-expression patterns of these identified metabolites, we conducted a clustering analysis applying K-nearest neighbor (KNN), shared nearest neighbor (SNN) and Louvain algorithms and revealed that the 527 measured metabolites could be classified into eight major clusters (**Fig. 1b-c, Additional file 1: Fig. 1 and Additional file 2: Table S2**). Functional enrichment analysis based on customized class and pathway terms was further performed for each cluster to identify shared pathways among groups of co-expressed metabolites (**Fig. 1d and Additional file 2: Table S3**). Specifically, cluster 1 showed a co-regulation of pathways involved in fatty acid metabolism, including steroids and their derivatives, as well as monoglycerides. Cluster 2 mainly included metabolites related to the metabolism of dietary components such as caffeine and benzoates. Cluster 3 included pathways central to the urea cycle, specifically the arginine and proline metabolism. It also involved the metabolism of amino acids like methionine, cysteine, glycine, serine, and theronine, which are crucial for nitrogen balance and protein turnover^61^. Cluster 4 exhibited a co-expression similar to Cluster 3, featured by metabolites like dipeptides as well as the metabolisms of arginine, aspartate, and gamma-glutamyl amino acids, which are mainly involved in protein synthesis and amino acid recycling^62^. Cluster 5 mainly consisted of metabolites essential for the cell membranes composition and signaling, including sphingomyelin, ceramide, and phosphatidylcholine^63^. Cluster 6 showed the interconnected pathways of glycolysis, gluconeogenesis, and sugar metabolisms, including the metabolism of fructose, mannose, and galactose. Cluster 7 included metabolites related to energy storage and mobilization, such as triglycerides and diglycerides^64^. Cluster 8 comprises lyso-phosphatidylcholine, lyso-phosphatidylethanolamine, and phosphatidylcholine, key components involved in cell membrane remodeling^65^.

### Individual and seasonal variations in plasma metabolome profiles

We assessed the variability in the expression of each metabolite over time by analyzing both inter-individual and intra-individual variations, calculated using the coefficient of variance (CV; **Fig. 2a and Additional file 2: Table S4**). Notably, our analysis revealed that the variability between individuals for each measured metabolite was greater than the variability observed within the same individual, with ratios of inter-individual to intra-individual CV ranging from 1.09 to 8.86. Among these metabolites, pyrrole-2-carboxylic acid and picolinic acid showed the most significant differences between individuals (**Fig. 2a**). This suggests that, despite the fluctuations in each person’s metabolic profile over time due to various environmental factors, each individual maintained a distinct metabolomic signature.

**Fig. 2.**
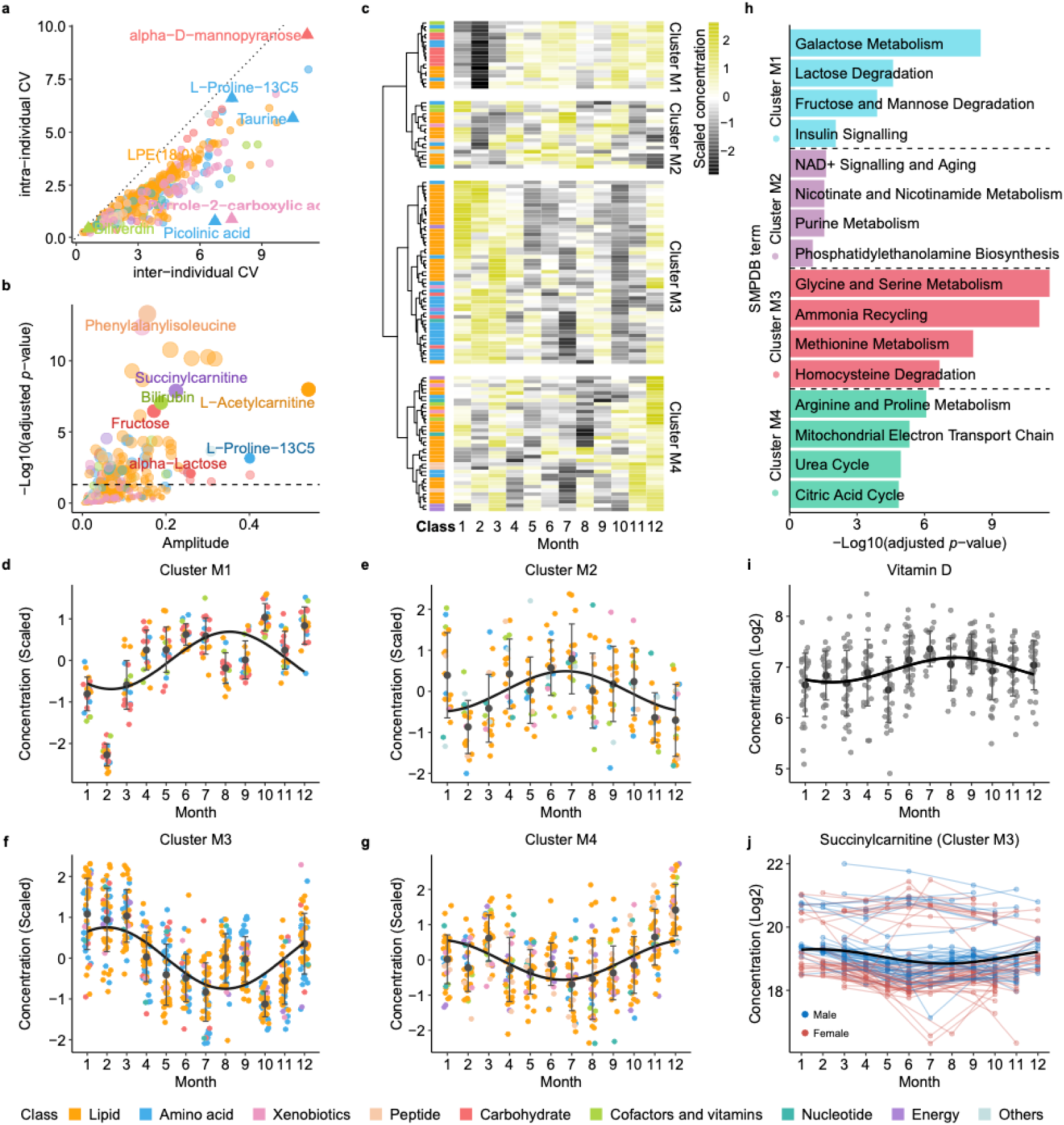
Inter- and intra-individual variability of plasma metabolites and seasonal influences. **a** The inter-individual and intra-individual variations of plasma metabolite levels, calculated as the coefficient of variation (CV) for each metabolite within each visit and across all participants, color-coded by metabolite classes. **b** Seasonal variation analysis of plasma metabolites using amplitude analysis, color-coded by metabolic classes. Y-axis showing the adjusted *p*-values with multiple test corrections using Benjamini and Hochberg method**. c** Heatmap showing the scaled levels of 121 metabolites with significant seasonal variations across 12 months. **d-g** Plasma metabolite levels throughout the year for metabolites in Cluster M1-M4. **h** Pathway enrichment analysis of metabolites within each seasonal cluster. Significantly enriched pathways were defined with adjusted *P*-values < 0.05 (Fisher’s exact test with multiple test corrections using using Benjamini and Hochberg method). **i**, Vitamin D concentration levels across 12 months during the study period. **j**, Succinylcarnitine levels over 12 months as an example of metabolites in Cluster M3. Each line represents an individual; red lines indicate females and blue lines indicate males. Regression lines are added using trigonometric functions.

Interestingly, some metabolites showed seasonal patterns which partially contributed to the intra-individual variations. To explore the seasonal influences on metabolite variability and identify potential seasonal patterns, we performed an association analysis between metabolite levels and the month of sampling, with sex, age and BMI considered as covariates. A total of 121 metabolites showed clear associations with the month of sampling, with amplitudes ranging from 0.016 to 0.541 (**Fig. 2b and Additional file 2: Table S5**). Hierarchical clustering of these metabolites further showed four distinct seasonal patterns: extremely low levels from January to March (Cluster M1, n = 18); relatively low levels during the winter months (Cluster M2, n = 18); relatively high levels during winter (Cluster M3, n = 49); extremely high levels in December (Cluster M4, n = 36) (**Fig. 2c-g and Additional file 2: Table S6**). The metabolites in Cluster M1, exhibiting the lowest levels from January to March, were enriched in carbohydrate metabolism pathways (**Fig. 2h and Additional file 2: Table S7**). As an example, fructose reached its lowest levels in January and February (**Additional file 1: Fig. 2a**). This could reflect a post-holiday shift to baseline dietary habits or a reduction in the consumption of sugars. Conversely, the elevation of energy-related pathways in Cluster M4 during December may indicate an increased caloric intake during the festive period or an adaptive physiological response to colder temperatures. Metabolites in Cluster M2, with relatively low levels in winter, were enriched in pathways associated with NAD^+^ signaling, nicotinate and nicotinamide metabolism, purine metabolism, and phosphatidylethanolamine biosynthesis (**Fig. 2h**). This could reflect a reduced requirement for or availability of these pathways’ end products during the winter, possibly due to changes in diet, reduced exposure to sunlight affecting vitamin D synthesis, which in turn affects NAD^+^ synthesis ^66^. Interestingly, the seasonal pattern of vitamin D closely correlated with the metabolites in Cluster M2 (Pearson *P* = 0.04827, **Fig. 2i**). In contrast, Cluster M3 metabolites, which were relatively high during winter, were associated with amino acid metabolism, specifically glycine and serine metabolism, as well as pathways involved in ammonia recycling and the metabolism of methionine and homocysteine (**Fig. 2h**). For instance, the highest level of cysteine was observed during winter (**Additional file 1: Fig. 2b**). These increased levels of amino acid metabolism could suggest a shift in fuel utilization towards amino acid catabolism for energy production and could also be associated with lower levels of physical activity as reported by Pedersen EB, *et al*^67^. Interestingly, the energy metabolism-related fatty acid succinylcarnitine in Cluster M3, which is involved in carnitine synthesis and utilization pathway, also showed a significantly high inter-individual variation, indicating varying levels of energy metabolism among individuals (**Fig. 2j**).

We further analyzed the seasonal variations of plasma proteins and found that some plasma proteins also exhibited seasonal patterns, with two opposite seasonal clusters observed: Cluster P1, with low levels during the colder season (Cluster P1, n = 51), and Cluster P2, with relatively low expression levels during the warmer months (Cluster P2, n = 312) (**Additional file 1: Fig. 2c-e and Additional file 2: Table S8**). As an example, 46 proteins in Cluster P2 were found to closely correlate with the seasonal patterns of metabolites in Cluster M3. These proteins primarily function in amino acid, glycan, and fatty acid metabolism. Specifically, angiopoietin-like protein 4 (*ANGPTL4*), which regulates lipoprotein lipase and has been found to be influenced by dietary fatty acids in both human muscle^68^ and mice heart^69^, showed associations with a group of fatty acids, such as cis-4-decenoylcarnitine and laurylcarnitine (**Additional file 1: Fig. 2f and 2g**). Along with these fatty acids, ANGPTL4 exhibited relatively lower expression levels from June to September (**Additional file 1: Fig. 2h**). The associations between proteins and metabolites will be described in more depth below.

### Sex- and BMI-related divergences in plasma metabolite levels

The associations between plasma metabolite concentrations, lifestyle factors and clinical measurements were analyzed using canonical correspondence analysis (CCA) that incorporated all 527 metabolites, 13 lifestyle variables, and 43 clinical chemistry and anthropometric variables across visits. Regression analysis of the two CCA components showed that CCA1 was mainly associated with BMI and lipid profiles, whereas CCA2 was more closely related to sex, body composition (muscle and fat percentage), hemoglobin, and urate. Notably, a clear divergence between male and female samples was observed, emphasizing sex as a significant factor influencing plasma metabolomic levels (**Fig. 3a**). The associations between metabolites and sex were visualized in a volcano plot (**Fig. 3b and Additional file 2: Table S9**). Among the 119 metabolites that showed significant sex differences, 35 were found to be elevated in females and 84 were more abundant in males (**Fig. 3b**). In particular, the peptide gamma-glutamylleucine exhibited higher levels in males (**Fig. 3c**) and has been found to be associated with elevated cardio-metabolic risks^70^. Additionally, 86 BMI-related metabolites were identified (**Fig. 3d and Additional file 2: Table S10**), with glutamic acid showing the strongest association (**Fig. 3e**), which aligns with previous reports^71^. Furthermore, the ratio of glutamic acid to other amino acids, such as lysine, ornithine, and hydroxyproline, have been reported as promising biomarkers for identifying metabolically healthy obese individuals^71^. Interestingly, lifestyle factors such as physical activity, stress, and smoking were found to correlate with metabolite levels and show collinearity with sex. In general, the stress levels in females were higher than in males, which was also significantly associated with the elevation of the stress hormone corticosteroid^72^. On the other hands, a higher incidence of smoking among males was associated with the alterations in several metabolites, such as glutamine, which plays a central role in cellular metabolism and function, highlighting the influence of lifestyle factors on metabolic health.

**Fig. 3.**
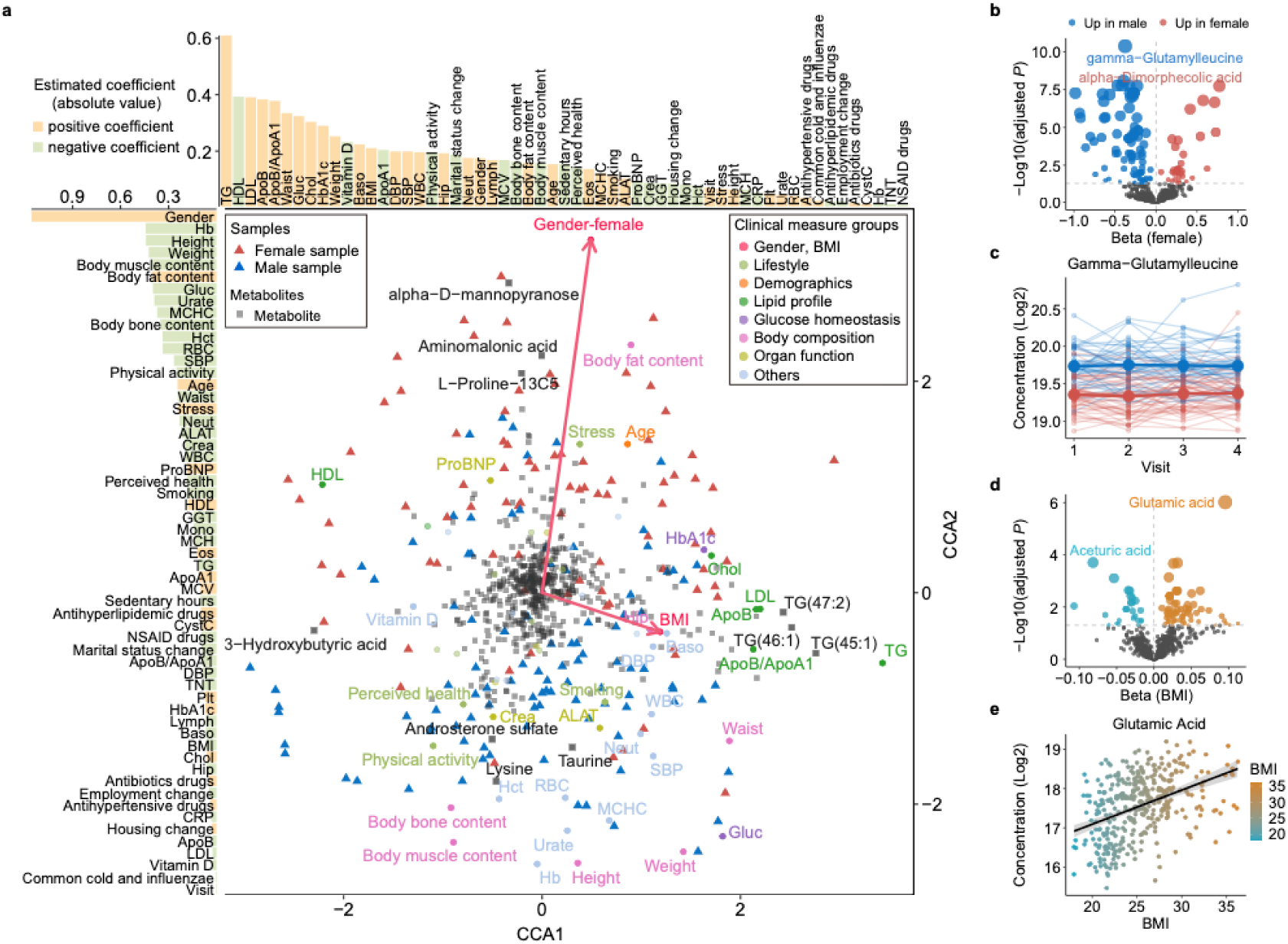
Influence of clinical measurements and lifestyle factors on metabolite levels. **a** Canonical correspondence analysis (CCA) showing correlations between plasma metabolite levels, clinical measurements and lifestyle variables. The upper and left bar plots show the estimated linear regression coefficients for clinical and lifestyle variables with respect to CCA1 and CCA2. **b, d** Volcano plots showing the impacts of sex (**b**) and BMI (**d**) on plasma metabolite levels (Kenward-Roger approximation with Benjamini and Hochberg correction). **c** Concentration of gamma-glutamylleucine across four study visits, shown as an example of a sex-associated metabolite. Each line represents an individual; red lines indicate females and blue lines indicate males. **e** Scatter plot showing the correlation between glutamic acid concentration and BMI, color-coded by BMI.

### Genome-wide association analysis of the plasma metabolite profile

To investigate the genetic influence on inter-individual differences in plasma metabolite concentration, a GWAS was performed based on individual variation coefficients for 527 plasma metabolites and 6.7 million common genetic variants (minor allele frequency, MAF > 0.05) identified through whole-genome sequencing. A total of 66 significant associations between genetic variants and individual blood metabolite concentrations were identified (*P* < 2.17 × 10^−9^, conventional *P* of 5 × 10^−8^ adjusted for the number of independent metabolites ^4^). Among them, 19 independent metabolite quantitative trait loci (mQTLs) (Linkage Disequilibrium, LD, *r*^2^ < 0.1, conditional *P* < 0.01) involving 22 metabolites were identified (**Fig. 4a and Additional file 2: Table S11**). Of 19 mQTLs, 4 were pleiotropic genetic variants associated with multiple metabolites. Of these metabolites, 45% were lipids (n = 10) (**Fig. 4a-b**). Out of the 22 genetically associated metabolites in our study, six have been previously analyzed in other GWAS studies^4,7^. To validate the associations between these metabolites and genetic variants, a meta-GWAS analysis was conducted for these six metabolites. Most of the genetic loci (8 out of 11) identified from meta-analysis showed the same direction of effects as in our study (**Additional file 2: Table S12**). Among these, the association between the genetic variant (rs34673751) from Acyl-CoA dehydrogenase short chain (*ACADS*) and butyrylcarnitine was the most significant in the meta-analysis. The *ACADS* gene encodes the enzyme short-chain acyl-CoA dehydrogenase (SCAD), which is essential for mitochondrial fatty acid beta-oxidation, while butyrylcarnitine is a short-chain acylcarnitine involved in fatty acid transport and energy metabolism. Our longitudinal analysis further demonstrated that individuals carrying A allele at rs34673751 exhibited stable higher blood butyrylcarnitine levels. Moreover, heterozygous individuals for the protein variant presented intermediate levels of blood butyrylcarnitine compared to the homozygous groups **(Fig. 4c-e**). To experimentally validate the association between *ACADS* and butyrylcarnitine, we knocked down the expression of *ACADS* in 293T cell lines using siRNA. Subsequent metabolite analysis revealed that silencing the *ACADS* gene increased the levels of butyrylcarnitine in these cells **(Fig. 4f-g**), providing direct evidence of *ACADS*’s role in the regulation of butyrylcarnitine levels.

**Fig. 4.**
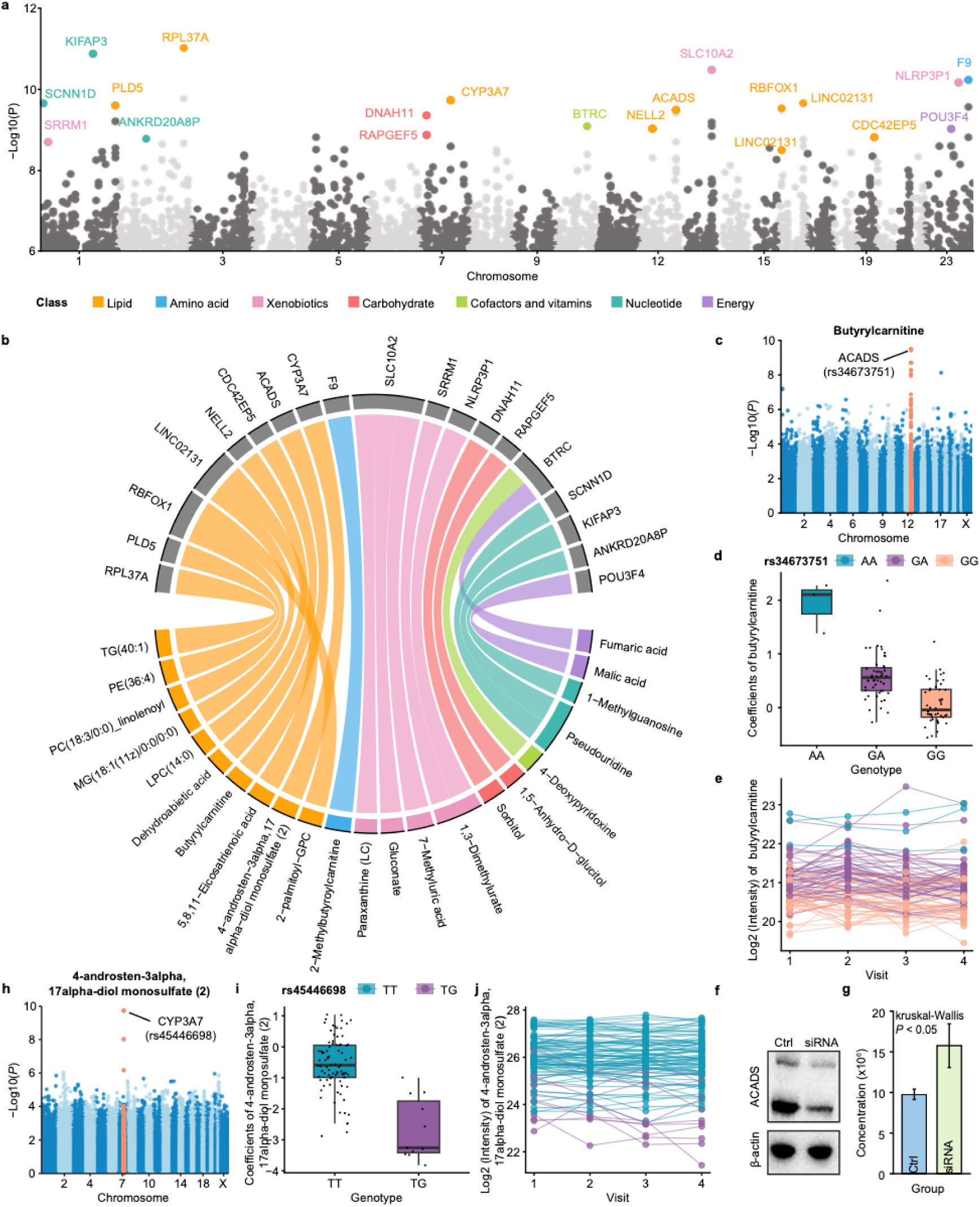
Genome-wide association analysis of the genetic regulation of the plasma metabolites. **a** Manhattan plot showing the identified mQTLs in the study. Significant loci are annotated based on the closest gene, with colors indicating the class of the corresponding metabolite. **b** Chord diagram of loci shared (*r*^2^ > 0.2) among metabolites in GWAS study. Line thickness is proportionate to the - Log10(*P*). **c** Manhattan plot of butyrylcarnitine showing the genome locations of all associated mQTLs. **d** Boxplot showing the association between plasma levels of butyrylcarnitine and the genotype of rs34673751, color-coded by the genotype of rs34673751. **e** Plasma levels of butyrylcarnitine across study visits; each individual is represented by a line; color-coded by the genotype of rs34673751. **f-g** Western blot showing increased butyrylcarnitine levels in the ACADS know-down 293T cell lines compared to the control group. **h** Manhattan plot for 4-androsten-3alpha,17alpha-diol monosulfate (2), showing the associated genetic loci. **i** Boxplot showing the association between plasma levels of 4-androsten-3alpha,17alpha-diol monosulfate (2) and the genotype of rs45446698. **j** Plasma levels of butyrylcarnitine across study visits; each individual is represented by a line; color-coded by the genotype of rs34673751.

Another notable example of the identified mQTLs is the association between metabolite 4-androsten-3alpha,17alpha-diol monosulfate (2), a sulfated steroid and a derivative of androstenediol, and the gene cytochrome P450 family 3 subfamily A member 7 (*CYP3A7*). (**Fig. 4h**). The highest association was found for the variant rs45446698, located upstream of the *CYP3A7* gene. Individuals carrying a TT homozygote had higher and more stable levels of 4-androsten-3alpha,17alpha-diol monosulfate (2) than individuals carrying a TG heterozygote (**Fig. 4i-j**).

### Quantification of genetic and non-genetic effects on plasma metabolite levels

To quantify the influence of genetics, lifestyle, and physiological conditions on metabolite variability, we applied a linear multivariate regression model to each metabolite. This model included all 19 mQTLs, 13 lifestyle factors, 43 anthropometric and clinical chemistry parameters, and visit. In the analysis, the genetic variants were combined as “genetic component”, all the lifestyle-related factors were combined as “lifestyle component”, and all anthropometric and clinical chemistry variables were categorized into 13 clinical classes. A summary of the analysis across all 527 analyzed plasma metabolites (**Fig. 5a and Additional file 2: Table S13**) showed that the influence of genetics, lifestyle, and physiological conditions on plasma metabolite level varied considerably. Genetics emerged as one of the important factors; out of the 22 metabolites with at least one significant genetic association, 5 metabolites had a genetic contribution greater than 20% (**Fig. 5b**). The metabolite most affected by genetics was 1,3-Dimethylurate, which is formed from caffeine and can be used as an indicator of caffeine metabolism ^73^, with 30% of its blood level variance explained by genetic variants. Besides genetics, 469 metabolites were influenced by various clinical factors, with a total contribution greater than 20%. Consistent with the CCA analysis (**Fig. 3a**), body composition and lipid profiles showed the most significant associations with plasma metabolite levels, with 69 metabolites associated with each of them. In addition, 54 metabolites were associated with urate levels, 48 with kidney function, 36 with glucose homeostasis, 18 with liver function, 17 with heart function, 15 with leukocytes, 6 with the acute phase response, and 28 with other clinical parameters. The top 25 most significant metabolites associated with clinical components were highlighted in **Fig. 5c**. As an example, a significant association was observed between body composition and pyroglutamylvaline, aligning with previous study that found pyroglutamylvaline to be positively associated with leg muscle^74^. Additionally, multiple metabolites have been identified as being associated with immune cell and red cell populations, as well as inflammatory biomarkers like C-reactive protein (CRP) (**Fig. 5c and Additional file 2: Table S13**). These results indicated the intricate crosstalk between metabolism, immune response, and hematopoiesis.

**Fig. 5.**
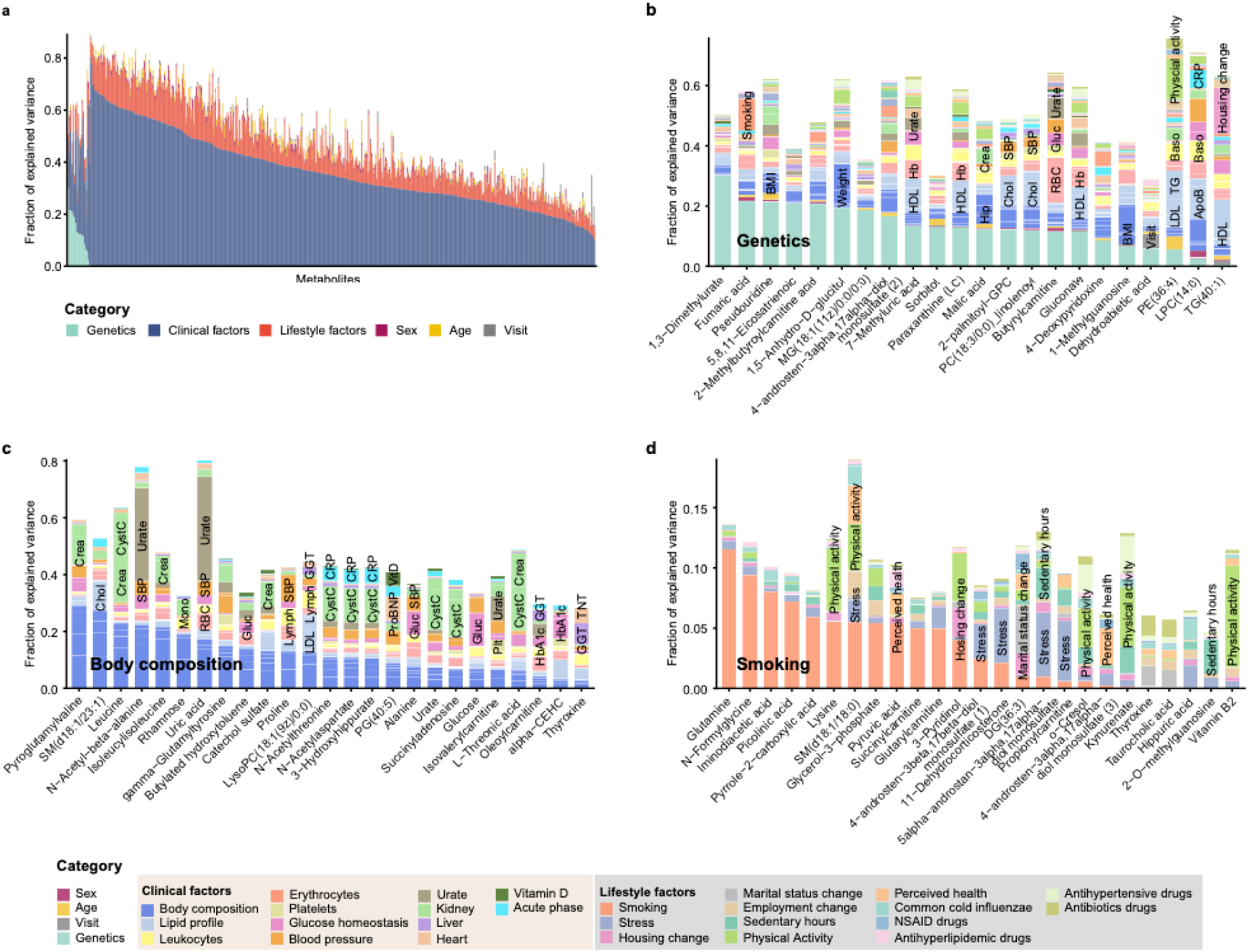
Influence of genetic, clinical and lifestyle factors on plasma metabolite level variability. **a** Overview of influence of genetic, clinical and lifestyle factors on the plasma metabolite level variability. **b** Barplot showing the variance explanation fraction for each component across all 22 genetic-related metabolites, color-coded by the variable classes. **c** Barplot showing the top 25 metabolites most strongly associated with clinical components. d Barplot showing the top 25 metabolites most strongly associated with lifestyle components.

Furthermore, we identified 39 metabolites associated with various lifestyle factors, with 9 showing an influence from lifestyle factors greater than 20% (**Additional file 2: Table S13**). These lifestyle-related metabolites included 15 associated with smoking, 8 with changes in housing, 4 with physical activity, 4 with stress, and 9 with other lifestyle factors. The top 25 most significant metabolites associated with lifestyle factors are listed in **Fig. 5d**. Notably, smoking had the most prominent influence on blood metabolite levels. Among the smoking associated metabolites, glutamine, the most abundant amino acid in the body and considered conditionally essential for critical illness and injury^75,76^, was negatively associated with smoking, along with factors such as 3-pyridinol, an intermediate product in nicotine degradation^77^ (**Additional file 2: Table S13**).

### Individual metabolite-protein profiles in human plasma

To investigate the co-expression patterns of plasma metabolites and proteins, we applied linear mixed modeling (LMM) to 527 metabolites and 794 proteins, adjusting for cofounders including subject, visit, sex, age, and BMI. The analysis revealed 5,649 significant protein-metabolite pairs, involving 479 metabolites and 625 proteins, each characterized by a correlation with a false discovery rate (FDR) adjusted *P* of less than 0.05 (**Additional file 2: Table S14**). Among these significant associations, 459 involving 121 metabolites and 257 proteins (48.93% of the overlapping metabolite-protein associations) aligned with previous findings from Benson MD, et al.^78^, despite differences in molecular measurement platforms. In **Fig. 6a** we present the 200 most significant protein-metabolite associations to illustrate the complex interplay within the protein-metabolite network. Multiple important hormones, enzymes, receptors and cytokines, such as glucagon (GCG), lipoprotein lipase (LPL), natriuretic peptide (NPPC), and interleukin 10 (IL10), have been identified as hub proteins in the network, highlighting their broad regulatory roles in human metabolism. Most of these protein-metabolite associations were connected to lifestyle and physiological conditions. As an example, we found a co-regulation between leptin (LEP) and aceturic acid **(Fig. 6b).** LEP, a hormone secreted by adipose tissue, plays an important role in regulating hunger and energy balance^79^, with higher concentrations observed in females^80^. Aceturic acid, also known as N-acetylglycine, is a derivative of the amino acid glycine and has been reported to modulate weight and associated adipose tissue immunity ^81^. Our findings suggested a significant interaction between LEP and aceturic acid with gender stratification.

**Fig. 6.**
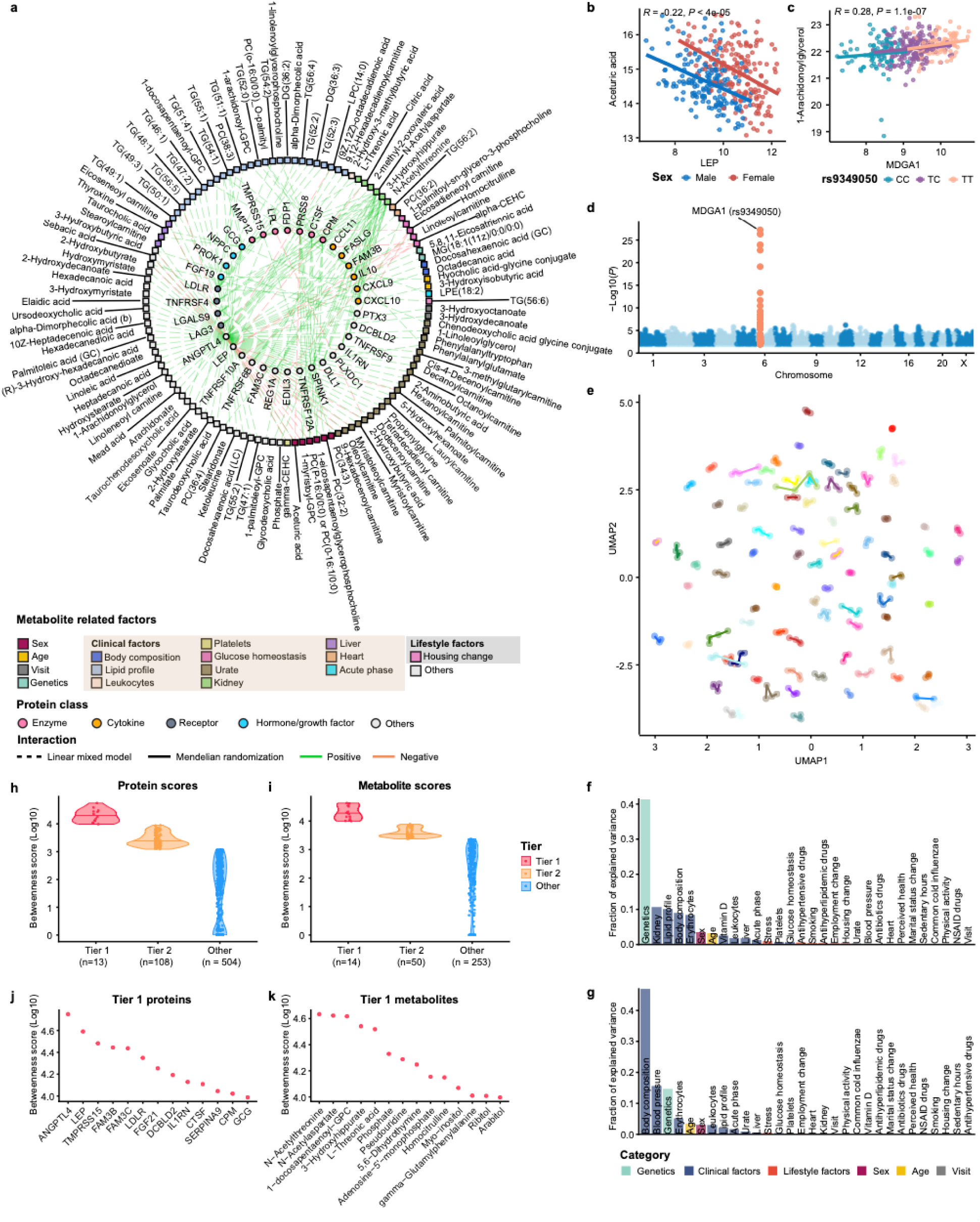
Characterization of protein-metabolite network. **a** Network presenting the top 200 significant protein-metabolite pairs identified by the linear mixed model (LMM) (FDR-adjusted *P* < 0.05). Solid circles represent proteins in the inner ring, color-coded by protein annotation. Squares represent metabolites in the outer ring, color-coded by different metabolite-related influence factors. Pairs of related proteins and metabolites are connected by dashed lines (indicating correlations supported by LMM results) and solid lines (indicating correlations supported by both the LMM and Mendelian randomization (MR) analysis). Green lines indicate positive correlations between proteins and metabolites in the LMM, while red lines indicate negative correlations. **b** Scatter plot showing the correlation between aceturic acid concentration and LEP, color-coded by sex. **c** Scatter plot showing the correlation between 1-Arachidonoylglycerol (1-AG) concentration and MDGA1, color-coded by different genotypes of rs9349050. **d** Manhattan plot for MDGA1 protein, showing the associated genetic variants with plasma levels of MDGA1. One of the most significant SNP (rs9349050) was used as an instrumental variable in the MR analysis. **e** UMAP clustering of the protein-metabolite profiles of the analyzed samples, color-coded by individual with lines connecting the visits for each individual. **f, g** Bar plots showing the variance explanation fraction of different genetic, clinical and lifestyle factors, calculated from linear mixed modeling, for UMAP1 (**f**) and UMAP2 (**g**). **h, i** Distribution of betweenness score of proteins (**h**) and metabolites (**i**) in three tiers. **j, k** Dot plots highlighting the Tier 1 proteins (**j**) and metabolites (**k**)

Subsequent Mendelian Randomization (MR) analyses were conducted to investigate potential genetic drivers of causality between plasma proteins and metabolites. A total of 87 putative causal associations were identified between 38 proteins and 61 metabolites (FDR-adjusted *P* < 0.05, **Additional file 1: Fig. 3a, Additional file 2: Table S15**). As an example, we detected a significant MR association between MDGA1, a cell surface glycoprotein involved in cell adhesion, migration, axon guidance, and neurodevelopment^82–84^, and 1-Arachidonoylglycerol (1-AG) (**Fig. 6c**), a stable regioisomer of the endocannabinoid 2-AG engaged in physiological functions such as emotion, cognition, and neuroinflammation^85,86^. Our analyses showed that the cis instrumental variable (rs9349050, **Fig. 6d**) for the MDGA1 protein consistently differentiated both MDGA1 and 1-AG levels in individuals carrying different genotypes (**Additional file 1: Fig. 3b-c**). This stable and positive association between MDGA1 and 1-AG indicated a potential genetic basis for the co-regulation of proteins and metabolites in the nervous system.

Using Uniform Manifold Approximation and Projection (UMAP), we clustered the individual molecular profiles based on the integrated metabolite-protein network. We noted that each individual possessed a unique and stable protein-metabolic profile (**Fig. 6e**). Regression analysis of the two UMAP components (**Fig. 6f-g**) revealed that UMAP1 was most significantly associated with genetic factors, followed by kidney function, lipid profile, body composition and erythrocytes. UMAP2 showed significant associations with body composition, blood pressure, genetics, and erythrocytes. Additionally, immune response, vitamin D, urate levels, liver functions, glucose homeostasis were moderately associated with the metabolite-protein profiles. Lifestyle factors such as stress, drug intake, smoking, and physical activity also exhibited minor influences on these profiles.

To identify key proteins and metabolites with the highest connectivity of the protein-metabolite network, we quantified their importance using the centrality betweenness score and categorized them into three tiers based on their deviation from the median level, measured in median absolute deviations (MAD). These categories include Tier 1, high importance (beyond two MADs); Tier 2, moderate importance (beyond one MAD); and Other: low importance (within one MAD). (**Fig 6 h-i**). Notably, ANGPTL4 and LEP were the top two proteins with the highest scores (**Fig. 6j**)*. ANGPTL4* has been reported as a key regulator in lipid metabolism, primarily by inhibiting the activity of lipoprotein lipase (*LPL)*^87^, and also involved in angiogenesis, vascular permeability, and inflammation processes. In our analysis, we revealed that ANGPTL4 was associated with a broad spectrum of metabolites, involving those related to lipid and glucose metabolism, tissue functions, and immune responses **(Additional file 1: Fig. 3d).** LEP, which is a well-known hormone, has been shown in recent studies to reflect systemic alterations of the human metabolome^88,89^. In our analysis, we observed that LEP was associated with a spectrum of metabolites related to blood pressure and glucose homeostasis, as well as stress **(Additional file 1: Fig. 3d)**. On the other hand, N-Acetylaspartate (NAA), one of the most abundant metabolites in the mammalian brain, and 3-Hydroxyhippurate, a microbial aromatic acid metabolite derived from dietary polyphenols and flavonoids found in normal human urine, were identified as high-importance metabolites through network analysis and found to be associated with kidney function in the study **(Fig. 6k, Additional file 2: Table S13**).

### Variability of protein-metabolite profiles and the associations with metabolic health

To investigate the associations between protein-metabolite profile variability and metabolic health, we stratified the analyzed samples into two risk groups (high-risk group and low-risk group) based on the clustering patterns of five key indicators of metabolic syndrome: high density lipoprotein (HDL), body mass index (BMI), systolic blood pressure (SBP), triglycerides (TG) and fasting glucose (Gluc)^90^. The high-risk group was characterized by increased levels of BMI, SBP, TG, and Gluc, alongside decreased levels of HDL, in comparison with the low-risk group **(Fig. 7a**). We then explored the differences in clinical chemistry measurements between these two groups. We found significant higher levels of two liver function biomarkers (alanine aminotransferase, ALAT; gamma glutamyltransferase, GGT), one kidney function biomarker (urate), one heart function biomarker (troponin T, TNT), and one immune biomarker (white blood cells count, WBC) in the high-risk group, indicating an increased susceptibility to cardiometabolic diseases **(Fig. 7b**).

**Fig. 7.**
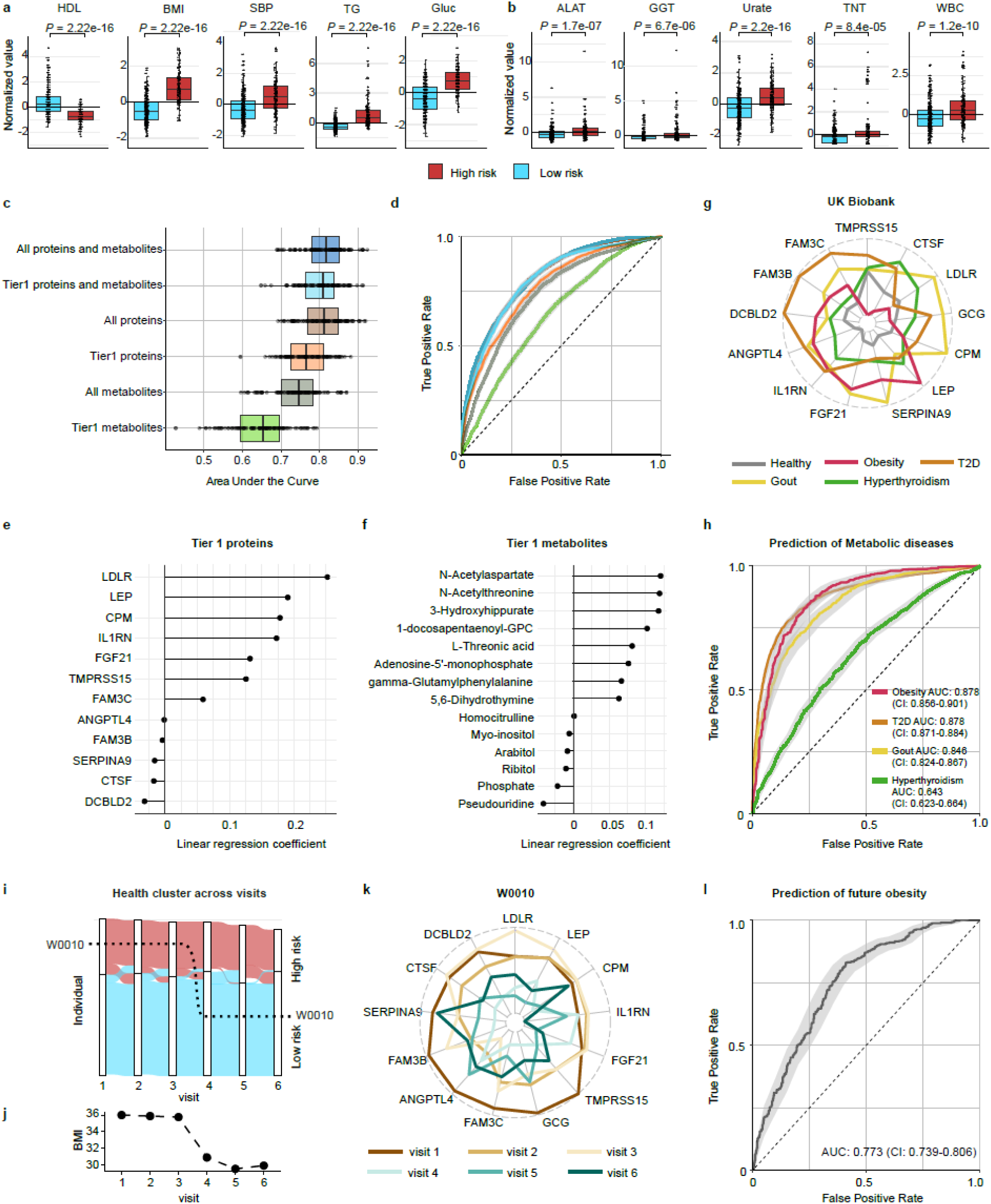
Predictive models for metabolic risk assessment. **a** Measurements of classical metabolic risk indicators, including HDL, BMI, SBP, TG and Gluc, compared between high risk and low risk groups. **b** Measurements of ALAT, GGT, urate, TNT and WBC for the high risk and low risk groups. **c** Area under the curve (AUC) of prediction models based on six different combinations of plasma proteins and metabolites: i) all proteins and metabolites; ii) tier 1 proteins and metabolites; iii) all proteins; iv) tier 1 proteins; v) all metabolites; vi) tier 1 metabolites. The AUC values were calculated by performing random sampling 100 times to account for variability. **d** Average receiver operating characteristic (ROC) curves for the predictive models for the six combination of plasma proteins and metabolites for predicting metabolic risk levels. **e, f** Linear model coefficients obtained using Tier 1 proteins (**e**) and metabolites (**f**) as independent variables and a dummy variable to indicate if the sample belonged to the high risk or ow risk group. **g** Tier 1 proteins’ profiles of healthy individuals, compared with obesity, T2D, gout or hyperthyroidism in the UK Biobank. **h** Average ROC curves obtained using Tier 1 proteins to predict individuals with either obesity, type 2 diabetes (T2D), gout, hyperthyroidism in the UK Biobank database. **i** Alluvial plot showing the risk level of individual samples across visits. The y-axis represents the individuals involved in the study, with colors indicating different risk levels. **j** BMI levels of the individual W0010 during the study visits. **k** Changes in Tier 1 proteins over time in individual W0010 who lost a significant amount of weight (15.4kg) between visit 3 and visit 4. **l** Average ROC curve of predictive models using Tier 1 proteins to predict future obesity in the UK Biobank. The 95% confidence intervals (CI) of ROC curves were plotted in **d, h, k** Red; High risk group, blue; Low risk group. High density lipoprotein, HDL; body mass index, BMI; systolic blood pressure, SBP; triglycerides, TG; fasting glucose, Gluc; alanine aminotransferase, ALAT; gamma glutamyltransferase, GGT, troponin T, TNT; white blood cells count, WBC.

Predictive modeling of the metabolic risk of the analyzed samples was conducted using proteins and metabolites. Interestingly, we found that the predictive power for risk groups, based on a combination of all proteins and metabolites, to be comparable to that based on Tier 1 proteins (n = 13) and metabolites (n = 14) alone, as well as models based on only proteins (**Fig. 7c,d**). This highlighted the potential importance of Tier 1 proteins and metabolites for metabolic risk assessments. In addition, proteins contributed more significantly to metabolic risk prediction than metabolites, with metabolites providing only marginal additional predictive value. Remarkably, the predictive power of Tier 1 proteins alone approached that of the full protein model (**Fig. 7c,d**), while metabolites demonstrated considerably lower predictive power compared to proteins (**Fig. 7c,d**). The importance of Tier 1 proteins and metabolites in relation to metabolic risk was quantified using a linear regression model to estimate the impact coefficients (**Fig. 7e,f**). We further examined the expressions levels of the 13 Tier 1 proteins in individuals with metabolic diseases, including obesity, type 2 diabetes (T2D), gout or hyperthyroidism, compared to healthy individuals using data from the UK Biobank^49^. Our analysis revealed that the expression levels of all Tier 1 proteins were significantly different (*P*<0.05) between healthy individuals and those diagnosed with any of these metabolic diseases **(Fig. 7g, Additional file 1: Fig. 4)**. Furthermore, the expression patterns of these 13 Tier 1 proteins varied significantly across different metabolic diseases, indicating their potential use as diagnostic biomarkers for various metabolic conditions. Moreover, we assessed the predictive power of Tier 1 proteins for these metabolic diseases, resulting in average AUCs ranging from 0.643 and 0.878 across different diseases (**Fig. 7h**).

Next, we examined the stability of the risk levels of participants throughout the two-year study. In general, the metabolic health status of most participants (70 out of 101) remained consistent, with 39 categorized within the high-risk group and 31 within the low-risk group at every visit (**Fig. 7i**). A notable exceptional case was participant W0010, who experienced a significant weight reduction from 120 kg during the third visit to 104.7 kg by the fourth visit (**Fig. 7j**). This remarkable change caused a shift from the high-risk group in the initial three visits to the low-risk group in the subsequent three assessments. Focusing on the Tier 1 proteins, we observed a significant decrease in their levels from the third to the fourth visit (**Fig.7k**). However, between the fourth and sixth visits, protein levels reversed despite no significant change in BMI (**Fig. 7k**), suggesting alterations in the individual’s underlying metabolism at the molecular level. Additionally, we found that nearly half of the Tier 1 proteins (6 / 13) displayed significantly different levels between lean (BMI < 25) and obese (BMI > 30) groups (**Additional file 1: Fig. 5**). We further predicted future obesity in individuals with a baseline BMI lower than 25 in the UK Biobank using the 13 Tier 1 protein expressions. The results showed that participants who later developed obesity (BMI > 30) already exhibited a distinct profile of Tier 1 proteins at baseline compared to those who maintained a BMI lower than 30 throughout the study period (**Additional file 1: Fig. 6a, b**). Predictive models demonstrated that baseline expression levels of Tier 1 proteins could predict future obesity with an average AUC of 0.773 with a 95% confidence interval of 0.739-0.806 (**Fig. 7l**).

## Discussion

In this study, we have performed a longitudinal multi-omics analysis on a group of clinically healthy participants aged 55 to 65 over two years to explore how genetics, lifestyle and physiological conditions affect individual metabolic profiles. We systematically examined the abundances and dynamics of 527 blood metabolites, which were categorized into nine major classes: lipids, amino acids, xenobiotics, peptides, carbohydrates, cofactors and vitamins, nucleotides, energy, and others. These metabolites were analyzed alongside their co-regulations with 794 proteins. By integrating whole-genome sequencing and extensive phenotyping data collected concurrently at each sampling time point, our results revealed an intricate interplay between genetic predispositions and environmental factors in shaping individual metabolic profiles.

One unique aspect of our study is the focus on the temporal dynamics of individual metabolic profiles. We identified eight co-expressed metabolite clusters based on longitudinal data and linked them to various metabolic pathways that are susceptible to both internal and external modulators, such as energy mobilization and dietary intake. By analyzing intra-individual metabolite variation over time, we observed four distinct seasonal patterns of plasma metabolites, with more fluctuations or extreme concentrations during the summer and winter. These seasonal fluctuations of metabolites might reflect changes in physical activity ^67^, diet ^91^, and metabolic rate ^92^, which are closely related to the season and might be regulated by seasonal gene expressions in multiple tissue types, including adipose tissue, brain, and gonadal tissue ^93,94^. Similar seasonal variations were observed in the proteomics profiling, suggesting a coordinated seasonal influence on both metabolite and protein levels.

In general, our analysis revealed higher inter-individual variations than intra-individual variations in plasma metabolite levels. By integrating whole-genome sequencing, we identified significant associations between genetic variants and blood metabolite levels (mQTLs). Numerous mQTLs have been identified within various populations^8–13,95^, and in this study, we used individual coefficients obtained from longitudinal data to better associate metabolite levels with genetic factors. Considering the significant seasonal variations observed in metabolite levels, this approach allows for more accurate quantification of individual metabolite levels for mQTL identification. Our findings indicated that many plasma metabolite levels during adult life were genetically predetermined at birth and remained stable within a range of dynamics under healthy conditions. Additionally, our results showed that lifestyle and clinical factors also significantly contributed to the variability in blood metabolite levels. Notably, 89% of (469 out of 527) metabolites exhibited at least a 20% variability attributable to lifestyle or clinical factors, indicating a substantial influence from lifestyle and physiological conditions on human metabolism. Among these, body composition and smoking were the top physiological and lifestyle factors influencing blood metabolite levels.

Proteins, as essential regulators and executors of metabolic processes, are critical components of human metabolic profiles, making their analysis essential for a comprehensive understanding of human metabolism. By integrating longitudinal metabolomics and proteomics data from the same individuals, we established a protein-metabolite co-expression network and found that each individual possessed a unique and stable protein-metabolic profile over time. This stability of individuals’ protein-metabolite profiles suggested that, despite the changes in environment, there is an inherent individuality to metabolic regulation. Our analysis showed that genetic factors were the most significant contributors to these profiles, with additional moderate influences from physiological conditions such as kidney function, blood pressure, and body compositions. Moreover, lifestyle factors like stress, smoking, and drug intake also had considerable impacts.

Furthermore, we identified key signature proteins and metabolites that characterize individual protein-metabolic profiles and assessed the predictive power of the most important Tier 1 proteins and metabolites for identifying individuals at high metabolic risk. Remarkably, the predictive performance of these Tier 1 biomarkers was comparable to that of the full set of proteins and metabolites, underscoring their potential utility in metabolic health monitoring. This finding also revealed a superior predictive power of proteins over metabolites. This is likely because proteins are directly involved in essential metabolic processes, function as signaling molecules and regulators, and generally exhibit greater stability and consistency over time. In our previous report, we also showed that plasma proteome exhibited most of the effects from clinical data^34,33^. We further validated this panel of 13 Tier 1 proteins in large-scale population data from the UK Biobank for the prediction of metabolic diseases. These biomarkers showed significant potential in diagnosing metabolic diseases. Although, all of the 13 proteins were elevated in various metabolic diseases, they exhibited different patterns in different metabolic abnormalities. Interestingly, we observed that the levels of the 13 proteins in an obese individual in the S3WP study initially decreased following weight loss but subsequently started to revert to the initial levels, despite a stable BMI during the study period. In the UK Biobank, we also noted that individuals who eventually became obese already exhibited an altered profile of Tier 1 proteins at a normal BMI and at baseline. These observations suggest that fluctuations in the proteome may precede BMI changes, highlighting their potential value in weight management strategies. However, it should be noted that more comprehensive analyses in large disease cohorts of these plasma proteins and metabolites must be performed to validate their use as clinical biomarkers. Another limitation of this study is the relatively small cohort size, which may not capture the full spectrum of individual metabolic variability across diverse populations. Thus, further validation and generalization of the findings in larger, more diverse cohorts and in clinical settings are necessary.

## Conclusions

In summary, our study provides a comprehensive longitudinal analysis on how genetics, lifestyle, and physiological factors influence the human metabolic profiles. Our results demonstrated the dynamic interplay between genetic predispositions and environmental factors, including seasonal variations, lifestyle, and physiological changes. By integrating proteomics and metabolomics data, we established a detailed protein-metabolite network and identified key molecular signatures that enhanced metabolic disease diagnostics and risk assessments. These findings offer promising avenues for improving metabolic health monitoring and developing future targeted interventions based on an individual’s unique metabolic profile.

## Data Availability

This study utilized participant-level datasets that have been securely deposited at the Swedish National Data Service (SND), a repository certified by the Core Trust Seal (URL: https://snd.gu.se/). The GC and LC-MS datasets are available as part of the full SW3P Wellness multi-omics dataset with the doi 10.5878/rdys-mz27. In compliance with patient consent and confidentiality agreements, these datasets are accessible for validation purposes only. Requests for access can be directed to SND via email. The evaluation of such requests will be conducted in accordance with relevant Swedish legislation. Additionally, for inquiries specifically on research within the scope of the S3WP program, interested parties are encouraged to contact the corresponding author directly. The code can be made available upon request to the corresponding author.

## Abbreviations

1-AG: 1-Arachidonoylglycerol
ACADS: Acyl-CoA dehydrogenase short chain
ALAT: Alanine aminotransferase
ANGPTL4: Angiopoietin-like protein 4
ANOVA: Analysis of Variance
ApoA1: Apolipoprotein A1
ApoB: Apolipoprotein B
AUC: Area under curve
BMI: Body mass index
CCA: Canonical correspondence analysis
CRP: C-reactive protein
CV: Coefficient of variance
CYP3A7: Cytochrome P450 family 3 subfamily A member 7
FDR: False discovery rate
GCG: Glucagon
GC-MS: Gas chromatography-mass spectrometry
GGT: Gamma glutamyltransferase
Gluc: Fasting glucose
GWAS: Genome-wide association studies
HDL: High density lipoprotein
HMDB: Human Metabolomics Database
IL10: Interleukin 10
IV: Instrumental variable
KNN: K-nearest neighbor
LC-MS: Liquid chromatography-mass spectrometry
LD: Linkage Disequilibrium
LDL: Low density lipoprotein
LEP: Leptin
LMM: Linear mixed modeling
LPL: Lipoprotein lipase
MAD: Median absolute deviation
MAF: Minor allele frequency
MDGA1: MAM domain containing glycosylphosphatidylinositol anchor 1
mQTL: Metabolite quantitative trait loci
MR: Mendelian randomization
NAD+: Nicotinamide adenine dinucleotide+
NPPC: Natriuretic peptide
NPX: Normalized Protein eXpression
NTproBNP: N-Terminal pro-brain natriuretic peptide
pQTL: Protein quantitative trait locus
QC: Quality control
Q-TOF: Quadrupole Time-of-Flight
ROC: Receiver operating characteristic
S3WP: Swedish SciLifeLab SCAPIS Wellness Profiling
SBP: Systolic blood pressure
SCAD: Short-chain acyl-CoA dehydrogenase
SCAPIS: Swedish CArdioPulmonary bioImage Study
siRNA: short interfering RNA
SMPDB: Small Molecule Pathway Database
SNN: Shared nearest neighbor
SNP: Single-nucleotide polymorphism
T2D: Type 2 diabetes
TG: Triglyceride
TNT: Troponin T
UMAP: Uniform manifold approximation and projection
WBC: White blood cells count

## Declarations

### Ethics approval and consent to participate

The Swedish SciLifeLab SCAPIS Wellness Profiling (S3WP) study has been approved by the Ethical Review Board of Göteborg, Sweden (registration number 407-15), and all participants provided written informed consent. The study protocol adheres to the ethical guidelines outlined in the 1975 Declaration of Helsinki.

### Consent for publication

Not applicable.

### Authors’ contributions

W.Z. conceived and designed the study. G.B. supplied clinical material. M.U., G.B., W.Z. and F.E., collected and contributed data to the study. W.Z., J.W., A.Z., and X.W. performed the data analysis. W.Z., J.W., A.Z., and X.W. drafted the manuscript. All authors read and approved the final manuscript.

## Acknowledgements

Original human sensitive data was processed under Project sens2018115. The integrative multi-omics analysis of the processed data was performed using computational resources provided by SNIC through the Uppsala Multidisciplinary Center for Advanced Computational Science (UPPMAX) under Project Berzelius-2024-200. Part of this research was conducted using the UK Biobank Resource under Application 99914. We thank the Plasma Profiling Facility at SciLifeLab in Stockholm for conducting the Olink analyses and Dr. Linn Fagerberg for data collection and data analysis in the S3WP program. We also thank the Proteomics and Metabolomics Platform at Guangzhou Laboratory for conducting the cell-line validation experiment using liquid chromatography-time-of-flight mass spectrometry (Q-TOF). We also express our gratitude to Yanfen Hong for her assistance with data acquisition and analysis.

## Additional file 1

Supplementary Methods and Figures. Additional methods on experimental validation and figures S1-S6.

**Fig. S1** Co-expression clustering of 527 analyzed metabolites.

**Fig. S2** Seasonal variation analysis of plasma metabolites and proteins.

**Fig. S3** Mendelian randomization protein-metabolite network.

**Fig. S4** Associations between Tier 1 proteins and metabolic diseases.

**Fig. S5** Association between Tier 1 proteins and obesity.

**Fig. S6** Prediction of future obesity in the UK Biobank.

## Additional file 2

Table S1-S15.

**Table S1.** Complete list of analyzed metabolites, proteins, and clinical parameters.

**Table S2.** Classification and co-expression patterns of 527 analyzed metabolites.

**Table S3.** Functional enrichment analysis of metabolite co-expression clusters.

**Table S4.** Inter- and intra-individual variations of 527 analyzed metabolites.

**Table S5.** Plasma metabolites with significant seasonal variation.

**Table S6.** Temporal co-expression patterns of seasonal associated metabolites

**Table S7.** Pathway enrichment analysis of metabolite co-expression clusters.

**Table S8.** Temporal co-expression patterns of seasonally associated proteins.

**Table S9.** Sex-associated plasma metabolites.

**Table S10.** BMI-associated plasma metabolites.

**Table S11.** Summary of identified independent mQTLs

**Table S12.** Meta-analysis of identified independent mQTLs.

**Table S13.** Contributions of genetics, lifestyle, and clinical factors to the variability of plasma metabolites.

**Table S14.** Comprehensive mixed-effect modeling analysis of protein-metabolite associations.

**Table S15.** Summary of one-sample mendelian randomization analysis.

