## Additional_file1_FigureS1-S6 for "Longitudinal analysis of genetic and environmental interplay in human metabolic profiles and the implication for metabolic health"

### Supplementary method

#### Experimental validation

293T cell line was cultured in DMEM (C11885500BT, gibco), supplemented with 10% fetal bovine serum (A5669701, gibco), 1x penicillin-streptomycin (E607011-0100, BBI), in a humidified atmosphere of 5% CO2 at 37ºC.

ACADS short interfering siRNA1, GGAGTTGTTTCCCATTGCA and negative control siRNA (Beijing Tsingke Biotech) were purchased from Beijing Tsingke Biotech Co., Ltd. siRNA was diluted to a concentration of 100 μM. Firstly, cells (7×105 per well) were seeded in 6-cm plates and incubated at 37°C for 24 h. A total of 1.6 μL siRNA (50 nM) was added into 200 μL jetPRIME® buffer, and 8 μL jetPRIME® reagent (101000046, Polyplus) was added to the mix. Next, the transfection mix was incubated for 10 min at room temperature. The transfection mix was added to the cells in 3 ml complete medium dropwise and incubated at 37°C for 6-8 h. After 6-8 h, the supernatant was removed and fresh complete medium was added, and the following experiments were performed at 48 h after transfection.

The supernatant of 2×106 cells were collected from cell lysis at 48 h after transfection. The protein concentration in the supernatant was determined by Pierce™ BCA Protein Assay Kit. Equal amount of lysis buffer was added to 4-20% FuturePAGETM (ET15420LGel, ACE) and gel electrophoresis was performed. The lysates in the gel were then transferred onto 0.2uM PVDF membranes (ISEQ00010, Merck) for 30 mins. After the transfer, the membrane was blocked with 5% skimmed milk powder and incubated for 30 min at room temperature. The membrane was washed with TBST, then incubated with relevant primary antibodies (ACADS, 16623-1-AP, proteintech; β-actin, 81115-1-RR, proteintech) for overnight at 4°C. The secondary antibody (Goat anti-Rabbit IgG (H+L) HRP, nitrogen, 31460) was added onto this membrane at a dilution of 1:10000 and incubated at room temperature for 2 h. The membrane was added the ECL chemiluminescent liquid (34580, Thermo Fisher) and then placed into the imaging system for exposure. The relative content of butyrylcarnitine from 2 million cells was detected by a liquid chromatography-quadrupole time-of-flight mass spectrometer (LC-QTOF MS, Agilent #1290-6546).

### Supplementary figures

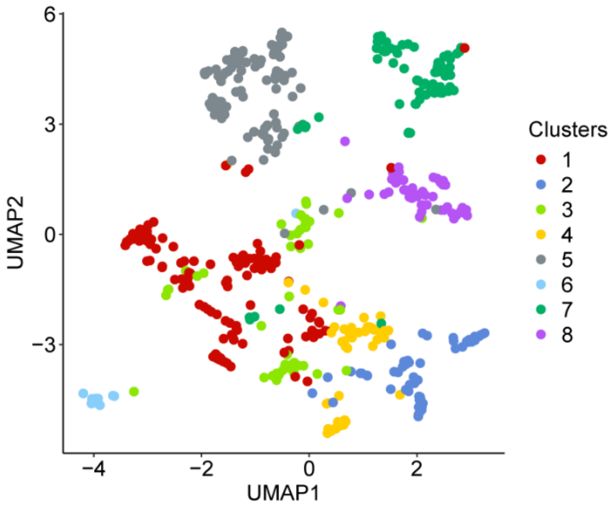


**Fig. S1 Co-expression clustering of 527 analyzed metabolites.**

UMAP clustering of the 527 metabolites measured in this study, showing their co-expression patterns, colored by clusters.

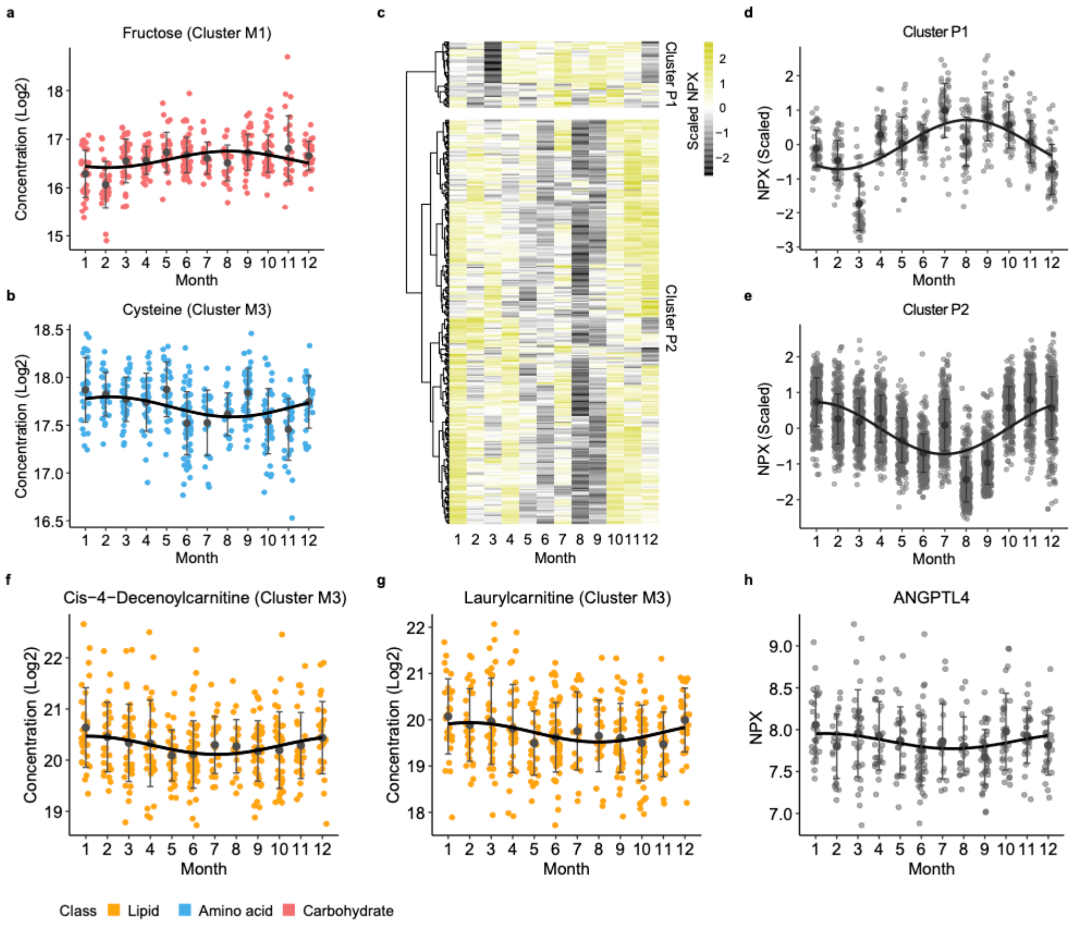


**Fig. S2 Seasonal variation analysis of plasma metabolites and proteins.**

**a, b, f, j** Scatter plots showing the plasma levels of fructose (**a**) and cysteine (**b**), and cis-4-decenoylcarnitine (**f**) and laurylcarnitine (**g**) across 12 months during the study period, color-coded by the metabolite classes. **c**, Heatmap showing plasma levels of 363 proteins with significant seasonal variations (adjusted $P < 0.05$, Kenward-Roger approximation with Benjamini-Hochberg correction). The plasma levels of proteins in each month were colored based on the scaled NPX values. **d, e** Scatter plots showing plasma metabolite levels throughout the year for proteins in Cluster P1 and P2. **h** Scatter plot showing the plasma levels of ANGPTL4 (angiopoietin-like protein 4) across 12 months. The regression curves were added in **a, b, d-h** by fitting trigonometric functions; and mean and standard deviation (SD) at each month were plotted.

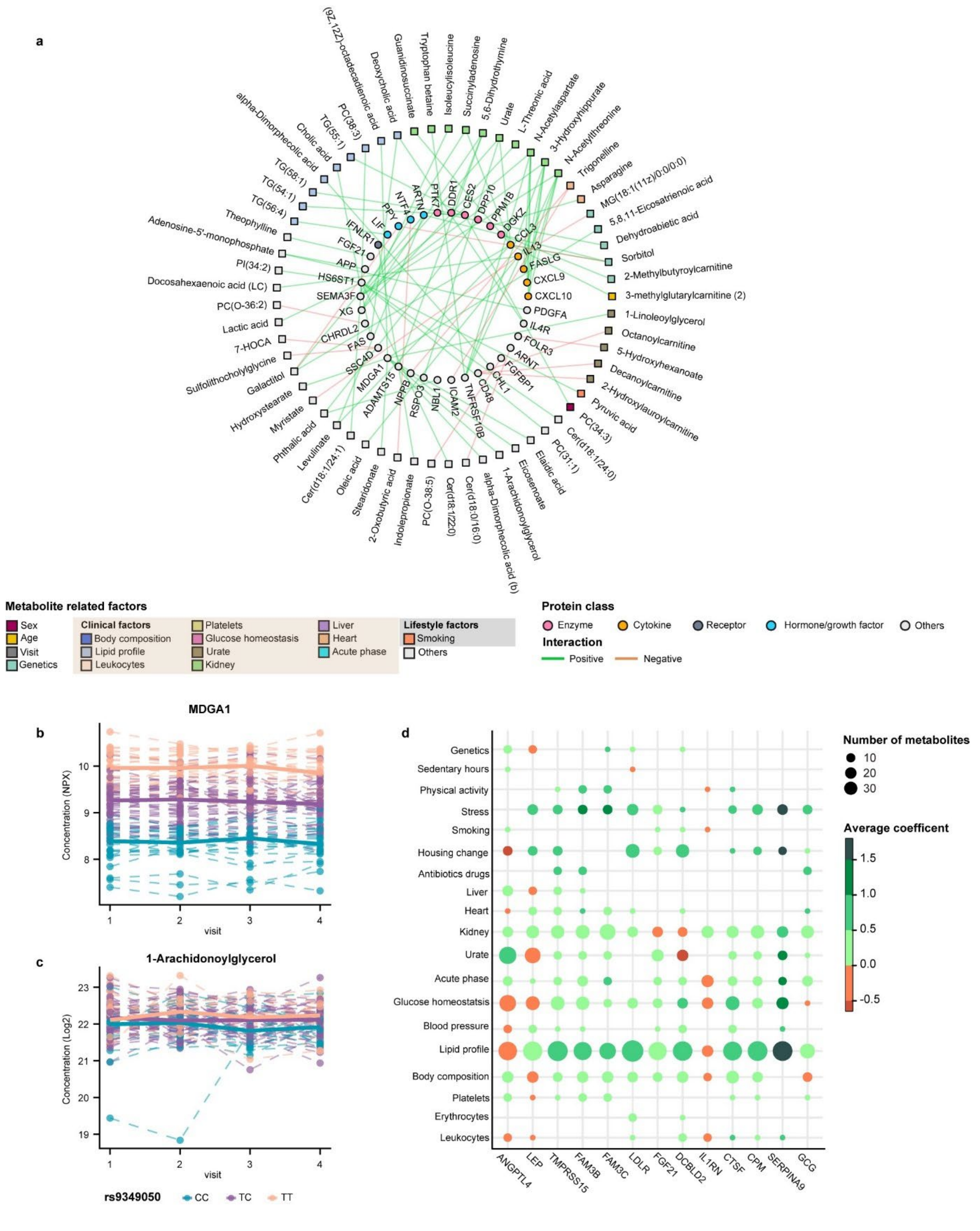
a
(9Z,12Z)-octadecadienoic acid
Deoxycholic acid
Guanidinosuccinate
Tryptophan betaine
Isoleucylisoleucine
Succinyladenosine
5,6-Dihydrothymine
Urate
L-Threonic acid
N-Acetylaspartate
3-Hydroxyhippurate
N-Acetylthreonine
Trigonelline
Asparagine
MG(18:1(11z)/0:0/0:0)
5,8,11-Eicosatrienoic acid
Dehydroabietic acid
Sorbitol
2-Methylbutyroylcarnitine
3-methylglutarylcarnitine (2)
1-Linoleoylglycerol
Octanoylcarnitine
5-Hydroxyhexanoate
Decanoylcarnitine
2-Hydroxylauroylcarnitine
Pyruvic acid
PC(34:3)
Cer(d18:1/24:0)
PC(31:1)
Elaidic acid
Eicosenoate
1-Arachidonoylglycerol
alpha-Dimorphecolic acid (b)
Cer(d18:0/16:0)
Cer(d18:1/22:0)
PC(O-38:5)
Indolepropionate
2-Oxobutyric acid
Stearidonate
Oleic acid
Cer(d18:1/24:1)
Levulinate
Phthalic acid
Myristate
Hydroxystearate
Galactitol
Sulfolithocholylglycine
7-HOCA
Lactic acid
PC(O-36:2)
Docosahexaenoic acid (LC)
PI(34:2)
Adenosine-5'-monophosphate
Theophylline
TG(56:4)
TG(54:1)
TG(58:1)
alpha-Dimorphecolic acid
Cholic acid
TG(55:1)
PC(38:3)
PTK7
DDR1
CES2
DPP10
PPM1B
DGKZ
CCL3
IL13
FASLG
CXCL9
CXCL10
PDGFA
IL4R
FOLR3
ARNT
FGFBP1
CHL1
CD48
TNFRSF10B
ICAM2
NBL1
RSPO3
NPPB
ADAMTS15
MDGA1
SSC4D
FAS
CHRDL2
XG
SEMA3F
HS6ST1
APP
FGF21
IFNLR1
LIF
PPY
NTF4
ARTN
Metabolite related factors
Sex
Age
Visit
Genetics
Clinical factors
Body composition
Lipid profile
Leukocytes
Platelets
Glucose homeostasis
Urate
Kidney
Liver
Heart
Acute phase
Lifestyle factors
Smoking
Others
Protein class
Enzyme
Cytokine
Receptor
Hormone/growth factor
Others
Interaction
Positive
Negative
b
MDGA1
Concentration (NPX)
visit
c
1-Arachidonoylglycerol
Concentration (Log2)
visit
rs9349050
CC
TC
TT
d
Genetics
Sedentary hours
Physical activity
Stress
Smoking
Housing change
Antibiotics drugs
Liver
Heart
Kidney
Urate
Acute phase
Glucose homeostatsis
Blood pressure
Lipid profile
Body composition
Platelets
Erythrocytes
Leukocytes
ANGPTL4
LEP
TMPRSS15
FAM3B
FAM3C
LDLR
FGF21
DCBLD2
IL1RN
CTSF
CPM
SERPINA9
GCG
Number of metabolites
10
20
30
Average coefficent
1.5
1.0
0.5
0.0
-0.5

**Fig. S3 Mendelian randomization protein-metabolite network.**

**a** Network representing all the significant (Benjamini Hochberg adjusted $P<0.05$) protein-metabolite interactions with genetic associations identified by Mendelian randomization (MR) analysis. Solid circles represent proteins in the inner ring, color-coded by protein annotation. Squares represent metabolites in the outer ring, color-coded by different metabolite-related variables. Pairs of related proteins and metabolites are connected by solid lines (indicating correlations supported by both the linear mixed model (LMM) and MR). Green lines indicate positive correlations between proteins and metabolites in the LMM, while red lines indicate negative correlations. **b, c** Longitudinal levels of MDGA1(**b**) and 1-Arachidonoylglycerol (1-AG) (**c**) across study visits, with each individual connected with a dash line. The median molecular levels were shown by the regression curves, color-coded by genetypes. **d** Bubble plot showing the numbers of metabolites associated with both Tier 1 proteins and genetic, clinical, and lifestyle variables. Color represents the average coefficients from the linear regression.

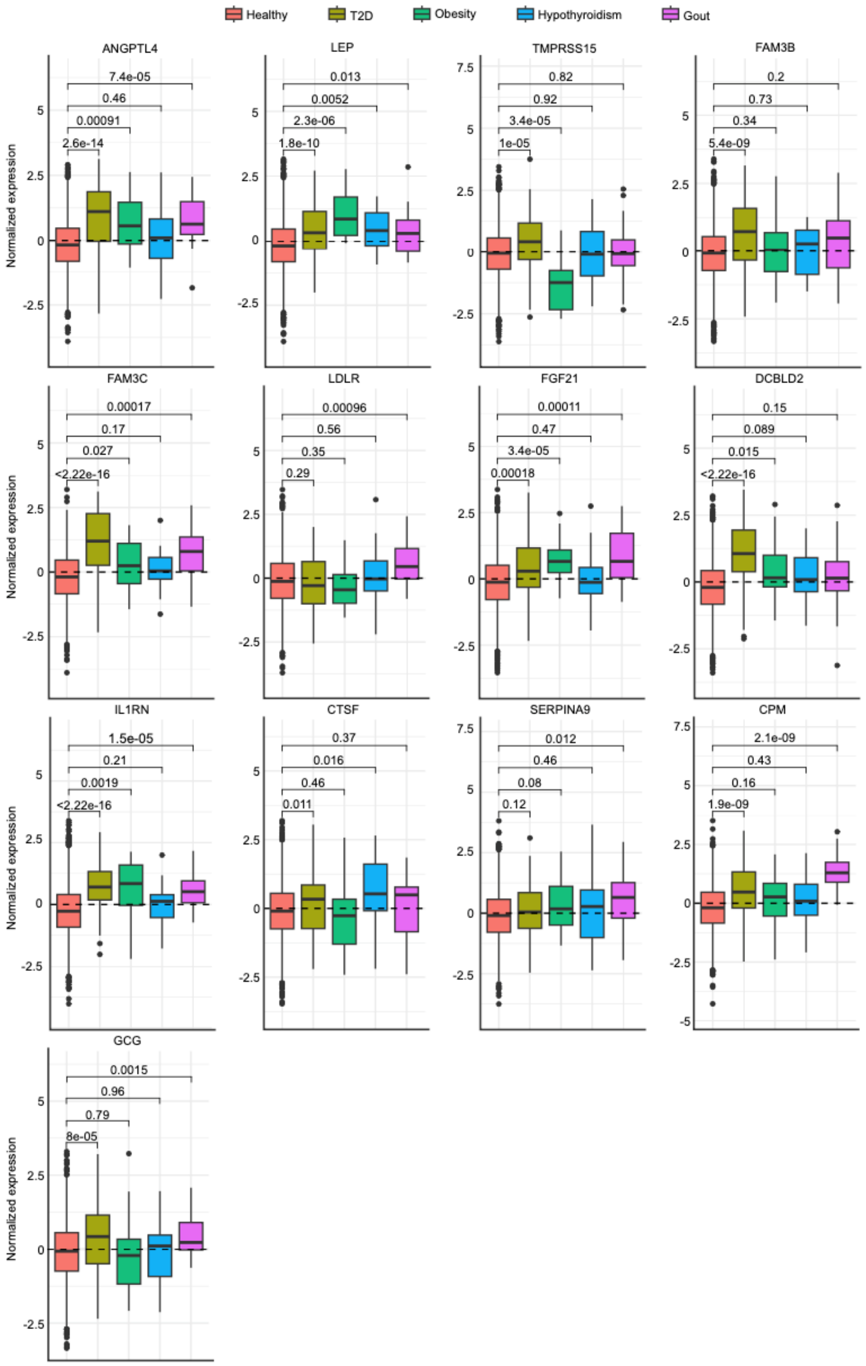


**Fig. S4 Associations between Tier 1 proteins and metabolic diseases.**

Boxplot showing the expression levels of Tier 1 proteins in individuals with metabolic diseases, including T2D, obesity, hypothyroidism, and gout, compared with healthy individuals, based on data from the UKBiobank.

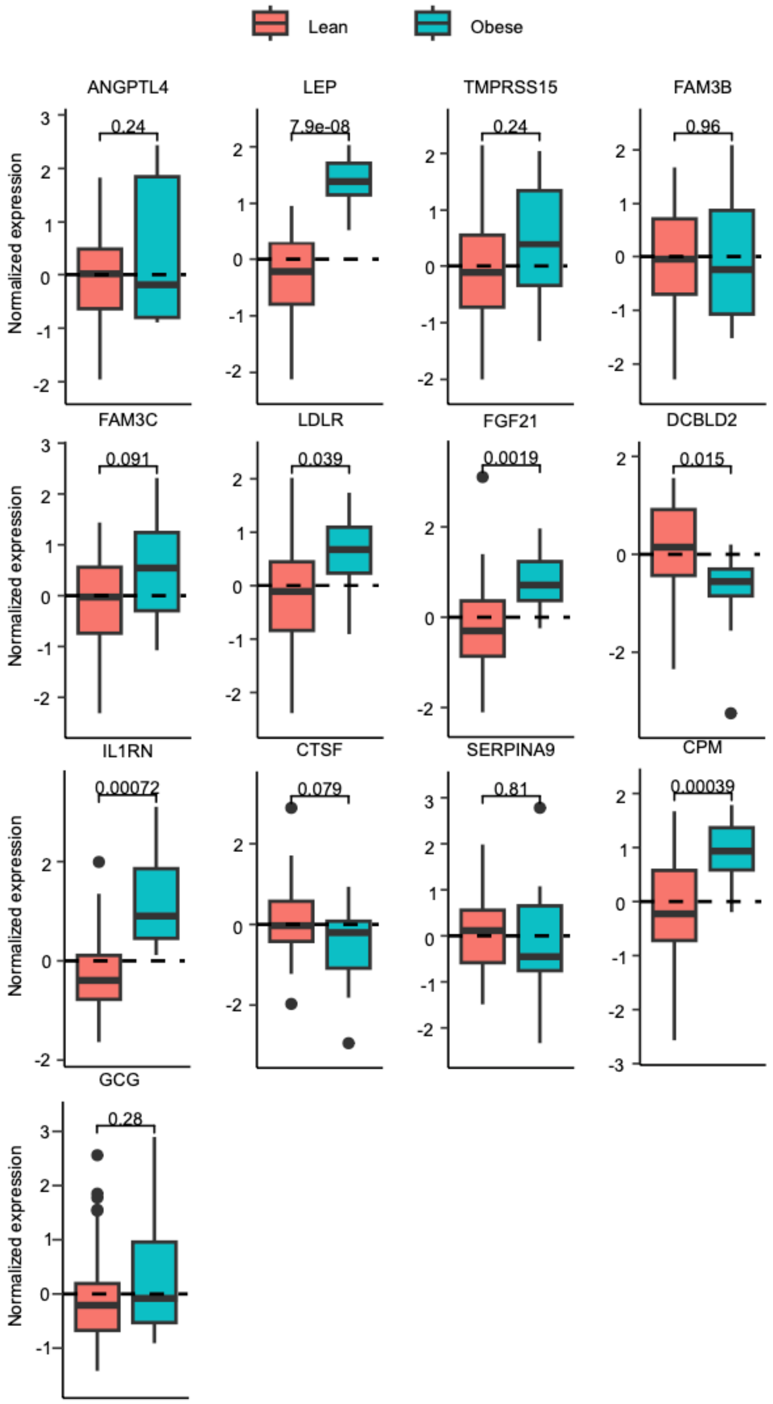


**Fig. S5 Association between Tier 1 proteins and obesity.**

Boxplot showing the expression levels of Tier 1 proteins in obese individuals (BMI > 30) compared to lean individuals (BMI < 25) based on data from the S3WP study.

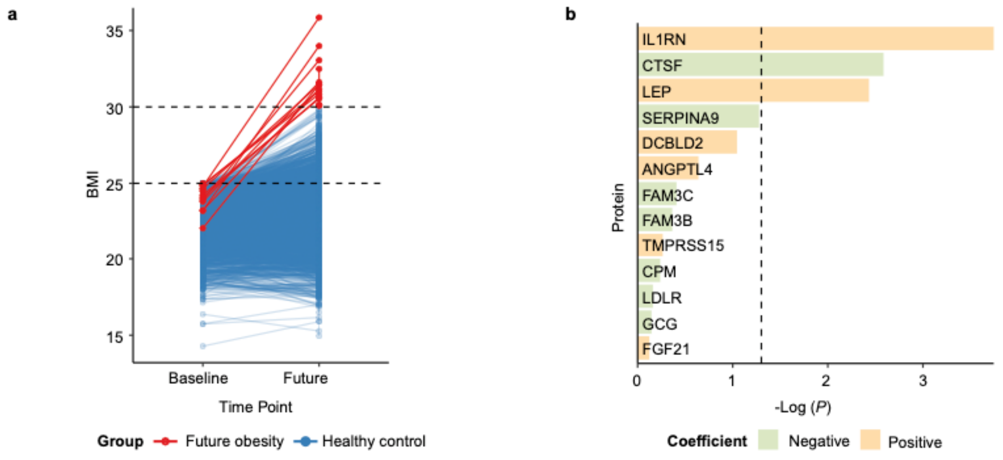


**Fig. S6 Prediction of future obesity in the UK Biobank.**

**a** Line plot showing the maximum BMI changes of individuals in the UK Biobank from baseline measurements during the study visits. Colors indicate whether an individual had a BMI > 30 at any of the subsequent visits. **b** Bar plot showing the coefficients of Tier 1 proteins for their associations with future obesity using a linear model. *P*-values were calculated by ANOVA with Benjamini-Hochberg correction for multiple testing.
